# Rare protein-disrupting variants in *NPY5R, DLGAP1* and *MAPK8IP3* segregate with OCD in two multiplex pedigrees

**DOI:** 10.64898/2026.04.21.26350600

**Authors:** Cathal Ormond, Mathieu Cap, Yi-Chieh Chang, Niamh Ryan, Denise Chavira, Kyle Williams, Jon E Grant, Carol Mathews, Elizabeth A Heron, Aiden Corvin

**Affiliations:** Neuropsychiatric Genetics Research Group, Department of Psychiatry, Trinity College Dublin, Dublin, Ireland; Department of Psychiatry, McKnight Brain Institute, Center for OCD, Anxiety, and Related Disorders, University of Florida, Gainesville, Florida, United States of America; University of Florida Genetics Institute, University of Florida, Gainesville, Florida, United States of America; Department of Psychology, University of California Los Angeles, Los Angeles, California, United States of America; Department of Psychiatry, Massachusetts General Hospital, Boston, Massachusetts, United States of America; Department of Psychiatry & Behavioral Neuroscience, University of Chicago, Chicago, Illinois, United States of America

**Keywords:** Obsessive compulsive disorder, pedigree, whole genome sequencing, rare variant, co-segregation, copy number variant

## Abstract

Obsessive compulsive disorder (OCD) is significantly heritable, but only a fraction of the contributory genetic variation has been identified, and the molecular etiology involved remains obscure. Identifying rare contributory variants of large effect would be an important milestone in helping to elucidate the mechanisms involved. Analysis of densely affected pedigrees is a potentially useful strategy to bypass the sample size challenges of standard case-control approaches. Here we performed whole genome sequencing (WGS) of 25 individuals across two multiplex OCD pedigrees. We prioritised rare variants using a Bayesian inference approach which incorporates variant pathogenicity and co-segregation with OCD. In the first pedigree, we identified a highly deleterious missense variant in *NPY5R*, carried by the majority of affected individuals. This gene is brain-expressed and has previously been implicated in panic disorder and internet addiction GWAS studies. In the second pedigree, we identified a large deletion of *DLGAP1* and a missense variant in *MAPK8IP3*, that perfectly co-segregated in a specific branch of the family: both genes have previously been implicated in OCD and autism. Both genes contribute to a protein interaction network including *ERBB4* and *RAPGEF1* which we had previously identified in a large Tourette Syndrome pedigree. Our analysis suggests that both energy homeostasis and downstream signalling from the post-synaptic density may both be important avenues for future research.

## 1 Introduction

Obsessive compulsive disorder (OCD) is a heritable psychiatric disorder with a global lifetime prevalence of approximately 1-2% (Ruscio, Stein, Chiu, & Kessler, 2010). The characteristic symptoms of recurrent obsessive thoughts and repetitive behaviours, typically emerge in middle childhood or early adulthood. The biology of OCD is poorly understood, prompting interest in identifying the molecular mechanisms involved. Large-scale consortium efforts have identified contributory common variants (Strom et al., 2025), and both rare copy number variants (M. W. Halvorsen et al., 2025), and protein-coding variants (Cappi et al., 2020; M. Halvorsen et al., 2021; Wang et al., 2025). However, collectively these only explain a fraction of heritability, and the necessity of having very large cohorts to detect true associations is a barrier to progress. As research groups turn to next-generation sequencing methodologies for primary analysis, this sample size requirement remains a major obstacle for gene-disease discovery. For rare variants, even those of large effect, observing a sufficient number of copies in a cohort may require tens of thousands of samples (Bose, Fuchsberger, & Boehnke, 2025; Lee, Abecasis, Boehnke, & Lin, 2014).

Family-based studies offer a complementary approach that typically requires smaller sample sizes. Given the proportion of DNA shared by close relatives, it is plausible that affected individuals in multiplex pedigrees are influenced by the same rare variant (Glahn et al., 2019). Family studies in OCD have highlighted complex inheritance patterns (Cavallini, Bertelli, Chiapparino, Riboldi, & Bellodi, 2000; Cavallini, Pasquale, Bellodi, & Smeraldi, 1999; Nestadt, Lan, et al., 2000), and while several linkage signals have been detected (Mathews et al., 2012; Ross et al., 2011; Willour et al., 2004), fine-mapping to genes of interest remains challenging. Nonetheless, first degree relatives of OCD cases have a higher risk for OCD (Blanco-Vieira et al., 2023; M. H. Huang et al., 2021; Nestadt, Samuels, et al., 2000) and related disorders such as Tourette syndrome (TS) (Mathews & Grados, 2011), hoarding disorder (Samuels et al., 2007) and body dysmorphic disorder (Altamura, Paluello, Mundo, Medda, & Mannu, 2001).OCD symptom dimensions are also correlated in multiplex pedigrees (Balachander et al., 2021). Indeed, family studies have already identified potential risk genes for traits related to OCD such as TS (Ercan-Sencicek et al., 2010; Ryan et al., 2022; Sun et al., 2018; Sundaram et al., 2011) and eating disorders (Bienvenu et al., 2019; Cui et al., 2013).

Here we examined two multiplex OCD pedigrees to identify rare variants likely implicated in phenotypic risk. Using whole genome sequencing (WGS), we evaluated a wide spectrum of gene-disrupting variants including single nucleotides (SNVs), indels, and copy number variants (CNVs). We also examined the contribution of common variants by generating polygenic risk scores (PRS) for OCD and related traits.

## 2 Methods

### 2.1 Sample recruitment and phenotyping

This study was approved by the Institutional Review Board at the University of Florida and ethical approval, contingent on local approval, was approved at Trinity College Dublin. As previously reported (Mathews et al., 2007), families were ascertained for ongoing genetic studies of OCD through probands who met criteria for DSM-5 OCD with onset in childhood (e.g., prior to age 18) and who did not have a diagnosis of pervasive developmental disorder, psychotic disorder, or bipolar disorder. Clinical assessments were conducted and DNA for genetic studies was obtained for all available members in each family, regardless of presumed diagnosis. Clinical assessments were conducted between 2002 to 2010 by psychiatrists or PhD level psychologists who were trained in research diagnostic assessments. Assessment instruments included the adult and child versions of the Yale Brown Obsessive Compulsive Scale (YBOCS (Goodman et al., 1989) and CYBOCS (Scahill et al., 1997) respectively), the Diagnostic Interview for Genetic Studies (DIGS) (Nurnberger et al., 1994), the Structured Clinical Interview for DSM-IV Axis I diagnoses (adults) (First, Spitzer, Gibbon, & Williams, 2012), the Schedule for Affective Disorders and Schizophrenia for School-Aged Children (Kiddie SADS) (children) (Kaufman et al., 1997; Orvaschel & Puig-Antich, 1987), and a semi-structured clinical interview that assessed for the presence and time-course of tics, stereotypies or other movement disorders (Pauls & Hurst, 1987). A best estimate consensus approach (described in (Mathews et al., 2012)) was used to assign all diagnoses. A diagnosis of definite OCD was given if the participant met all DSM-IV criteria for OCD. A diagnosis of probable OCD was given if the participant had multiple obsessions and compulsions, and some evidence of impairment, but did not meet full criteria for OCD (e.g., had mild symptoms that took less than an hour, impairment was denied, but symptoms caused some distress). The IDs of all individuals were only known to members of the research groups, so cannot be used for re-identification. For privacy regions, the sex of all individuals in the pedigree diagrams has been removed.

### 2.2 WGS data and quality control

We selected 25 samples across the two pedigrees for WGS – 15 samples from pedigree P1 and 10 samples from pedigree P2 (see Supplementary Table 1). We followed the previously described approaches for data pre-processing, quality control, and short variant calling for pedigrees (Ormond et al., 2023). We evaluated the pedigree consistency using peddy (Pedersen & Quinlan, 2017). We compared the genomic relatedness and the pedigree structure and resolved any discrepancies (see Supplementary Methods). For SNVs and indels, multi-allelic sites were normalised into multiple bi-allelic sites, and variants private to each pedigree were identified with SnpEff (Cingolani et al., 2012). Family-private variants were annotated with VEP (McLaren et al., 2016), with full details described previously (Ormond, Ryan, Cap, et al., 2024). Since all samples clustered with the EUR subset of the 1000 Genomes Project based on the peddy analyses (see Supplementary Figure 1), we took the allele frequencies from the non-Finnish European subset of the gnomAD v4.1 joint data (Karczewski et al., 2020).

CNVs were called using PECAN and were aggregated independently for both pedigrees (Ormond, Ryan, Byerley, Heron, & Corvin, 2024). CNVs carried by only one individual were removed before the main prioritisation analysis. Following previous evidence of an enrichment of rare CNVs in OCD (M. W. Halvorsen et al., 2025), we retained CNVs of length at least 30kbp that overlapped gene regions. Prioritised CNVs were manually validated by examining the sequencing reads of the surrounding region with samplot (Belyeu et al., 2021).

### 2.3 Variant prioritisation

BICEP was used to prioritise all rare variants (Ormond, Ryan, Cap, et al., 2024; Ormond, Ryan, Corvin, & Heron, 2026). BICEP quantifies how likely the variant is to be causal (logPostOC score) based on aggregated population-level metrics such as allele frequency, deleteriousness and genomic functional consequence (logPriorOC score), as well as the co-segregation with the trait of interest (logBF score). Individuals were considered a case if they had a confirmed or probable diagnosis of OCD. Where no diagnosis information was available, an individual was assumed to be a control.

In pedigree P1, all cases were descendants of individuals 141 and 142 (i.e., there were no affected married-in samples), and the inheritance pattern of OCD is broadly dominant. In pedigree P2, however, there were multiple unrelated individuals who are potential founders, of whom we prioritised two main founders: individual A and individual E (see Supplementary Figure 2). We therefore subdivided the output of BICEP to variants carried by individual A (an ancestor of seven of the eleven cases), and to variants carried by individual E (an ancestor of four of the eleven cases). We note that there is overlap between these two branches, so to avoid re-using data across analyses, we focused on variants that were private to each branch. When running BICEP on a branch, individuals from the other branch were removed from the pedigree structure, so as not to inappropriately adjust the co-segregation scores by including individuals who could never carry these private variants. Married-in individuals were not removed. In the output, we considered variants which were carried (or assumed to be carried) by the founder of the respective branch.

Previous work has shown an enrichment of rare *de novo* missense variants with high PolyPhen2 Scores in OCD cases compared to controls (M. Halvorsen et al., 2021; Wang et al., 2025). However, we found that the predictive ability of the BICEP prior model for missense variants performed noticeably poorer when we restricted the deleteriousness metrics to PolyPhen2 alone compared to the full model used in the original BICEP publication (see Supplementary Methods and Supplementary Figure 3). For this reason, we opted to use the original five deleteriousness metrics, which includes PolyPhen2.

### 2.4 Known rare risk variants and genes

We screened for known risk variants for OCD in all samples, regardless of the co-segregation status. For SNVs and indels, we examined 36 genes where ultra-rare protein-coding variants confer risk for OCD or chronic tic disorder (Wang et al., 2025). Following Wang et al., we filtered to variants that were ultra-rare in gnomAD (AF < 0.0005), and were either likely gene-disrupting (frameshift, stop-gain or splice donor/acceptor), or missense variants predicted to be deleterious (PolyPhen2 > 0.908).

To date, no individual CNV has been shown to be significantly associated with OCD (M. W. Halvorsen et al., 2025). However, CNVs associated with other psychiatric disorders, in particular autism and schizophrenia, have been observed to have pleiotropic effects (Rees & Kirov, 2021). We therefore compiled a list of variants (see Supplementary Table 2) associated with TS (A. Y. Huang et al., 2017), schizophrenia (Marshall et al., 2017; Rees, Walters, Chambert, et al., 2014; Rees, Walters, Georgieva, et al., 2014), autism (Sanders, 2015), and those identified from a cross-disorder analysis of six psychiatric disorders (Shanta et al., 2025). This list includes the 16p13.11 deletion that has been reported as suggestive for OCD (McGrath et al., 2014). The overlap criteria and positions were taken from Kendall et al. (Kendall et al., 2019) (with positions lifted from the original GRCh37 to GRCh38 with liftOver (Haeussler et al., 2019)) or from the cross disorder analysis. For single-gene CNVs, we typically required overlapping deletions to be exonic and overlapping duplications to cover the entire gene, both based on the MANE transcript (Morales et al., 2022).

### 2.5 Protein interaction network

We used the STRING database v12.0 (Szklarczyk et al., 2023) to visualise protein-protein interactions. As an input, we used all genes identified from the rare variant analysis in this study as well as four protein-coding genes (*NASP*, *RAPGEF1*, *ERBB4*, and *IKZF2*) identified from a previous family-based rare-variant analysis of TS performed by our group (Ryan et al., 2022). We removed the “textmining” source for the protein interactions, required a minimum interaction score of 0.9 (“highest confidence”), and selected 20 interactions in the first shell and zero in the second shell.

### 2.6 Principal components analysis

We performed a principal components analysis (PCA) of the pedigree members using the EUR subset of the 1000 Genomes Project samples (Auton et al., 2015) as a population reference. We obtained high-coverage WGS data for these samples from the EMBL-EBI FTP website (see Supplementary Methods, Web resources). We subsetted the pedigree and reference data to a list of shared variants and merged the two datasets. We removed variants in the MHC region, the known chromosome 8 inversion, as well as regions of high linkage disequilibrium (LD). Ambiguous SNPs were also removed, as were variants with <5% minor allele frequency, variants with >2% missingness or Hardy-Weinburg p-values less than 0.001.

Since a PCA is typically performed on unrelated samples, we first identified a maximally unrelated subset of individuals in our merged data, ran the PCA on the unrelated samples, and projected the remaining samples onto the principal component space using the “bigsnpr” package (Privé, Aschard, Ziyatdinov, & Blum, 2018) from R. Samples were considered unrelated if their pairwise kinship coefficients using the KING algorithm (Manichaikul et al., 2010) were less than 2^-4.5^ ≈ 0.044. This ensures that individuals are at least fourth-degree relatives (e.g., more distantly related than first cousins).

### 2.7 Polygenic risk scores

We obtained summary statistics from the Psychiatric Genomics Consortium website (see Supplementary Methods, Web resources) for the most recent genome-wide association study (GWAS) of OCD (Strom et al., 2025) to generate PRS. We also obtained summary statistics for TS (Yu et al., 2019), and panic disorder (PD) (Forstner et al., 2021), as these diagnoses have been observed in the pedigree members (see Supplementary Table 1) and have a shared common variant signal with OCD (Grotzinger et al., 2026). To compare the scores of the pedigree members, we used the EUR subset of the 1000 Genomes Project to generate a population-representative distribution of PRS. Since the summary statistics were supplied on genome reference build GRCh37, we converted our WGS data to this build using liftOver (Haeussler et al., 2019), first removing variants at known conversion-unstable positions (Ormond, Ryan, Corvin, & Heron, 2021).

The effect sizes from the summary statistics were adjusted with PRS-CS-auto (Ge, Chen, Ni, Feng, & Smoller, 2019), using the EUR subset of the UK Biobank as an LD reference panel and calculating the effective sample sizes from the original GWAS. PRS were then generated using plink (Purcell et al., 2007). Given that the cohort is not composed of independent samples, we used SAIGE (Zhou et al., 2018) to account for covariates as well as pedigree structure. First, a genomic relatedness matrix was generated on a LD-pruned subset of the variants, again taking individuals that are at least fourth degree relatives as unrelated. All scores were then adjusted for relatedness, sex, sequencing batch (i.e., reference or pedigree cohort), and the first 20 principal components by generating a null model with SAIGE. Finally, all scores were normalised to the EUR reference samples.

## 3 Results

### 3.1 Rare variants

#### 3.1.1 P1 pedigree

In the P1 pedigree, no sample contained a rare SNV or indel in any of the known 36 risk genes associated with OCD. From the BICEP analysis, no rare SNV or indel with a positive logPriorOC score had perfect co-segregation with OCD. Of the top 50 SNV/indel variants ranked by BICEP, the variant that ranked second had the best co-segregation with OCD and a positive logPriorOC score (see Figure 1A and Supplementary Table 3). This was an ultra-rare, highly deleterious missense variant in *NPY5R*, carried by 10 out of the 15 cases in the pedigree (see Figure 1B).

**Figure 1:**
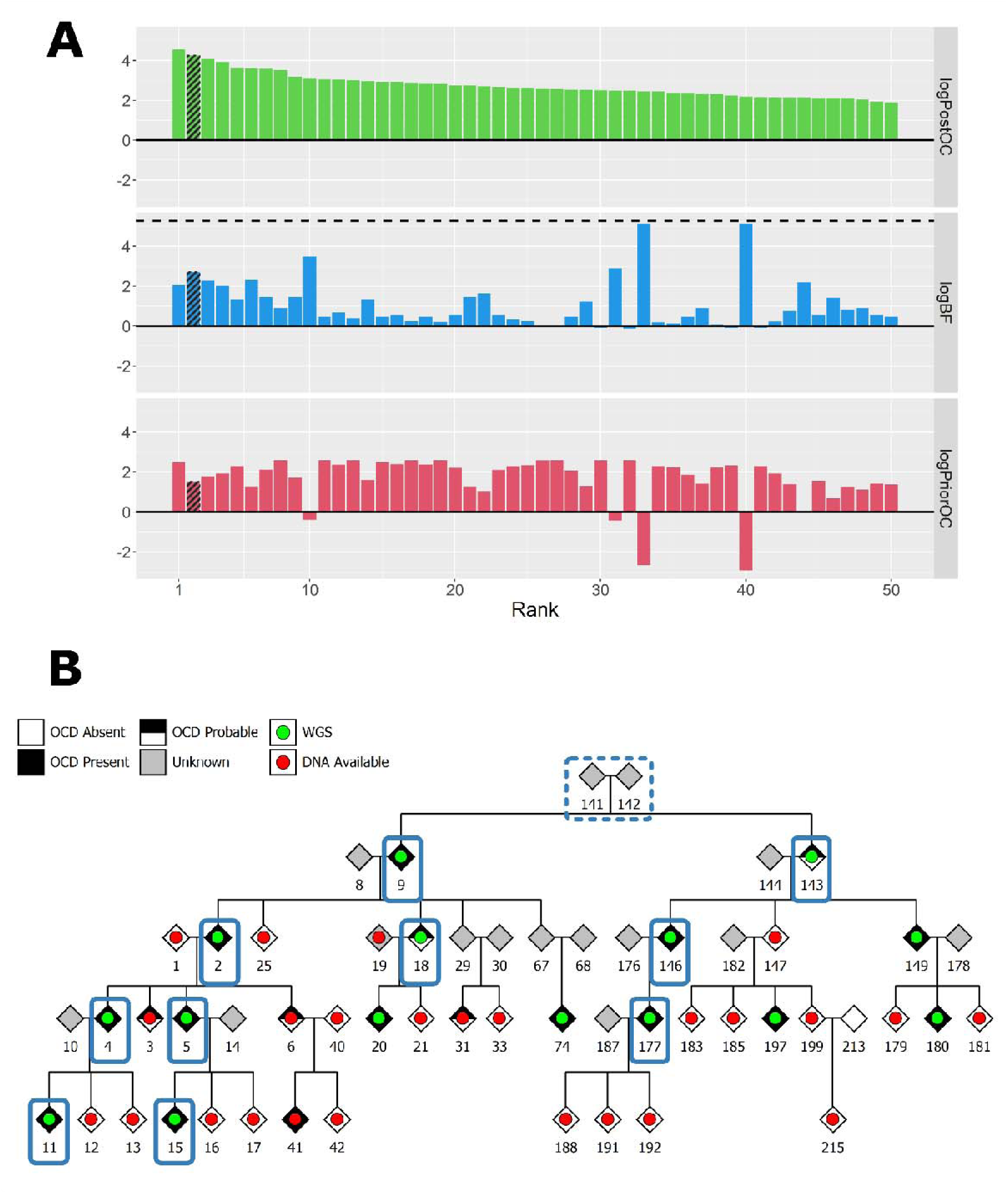
(**A**) BICEP output metrics for the top 50 ranked SNVs and indels in P1. The dashed line in the middle panel indicates the maximum logBF achievable in the pedigree. The shaded bars represent the *NPY5R* variant. (**B**) Pedigree diagram for P1. The blue rectangles indicate carriers of the *NPY5R* variant. Solid borders represent observed variant carriers, and dashed borders represent obligate carriers.

Our CNV calling and quality control resulted in 31 large, gene-disrupting variants (see Supplementary Table 4). However, all of these had either negative logPriorOC scores or poor logBF scores (see Figure 2). Additionally, we did not observe any of the established rare risk CNVs associated with the selected psychiatric disorders. Therefore, we do not see evidence of CNVs impacting OCD risk in this pedigree.

**Figure 2:**
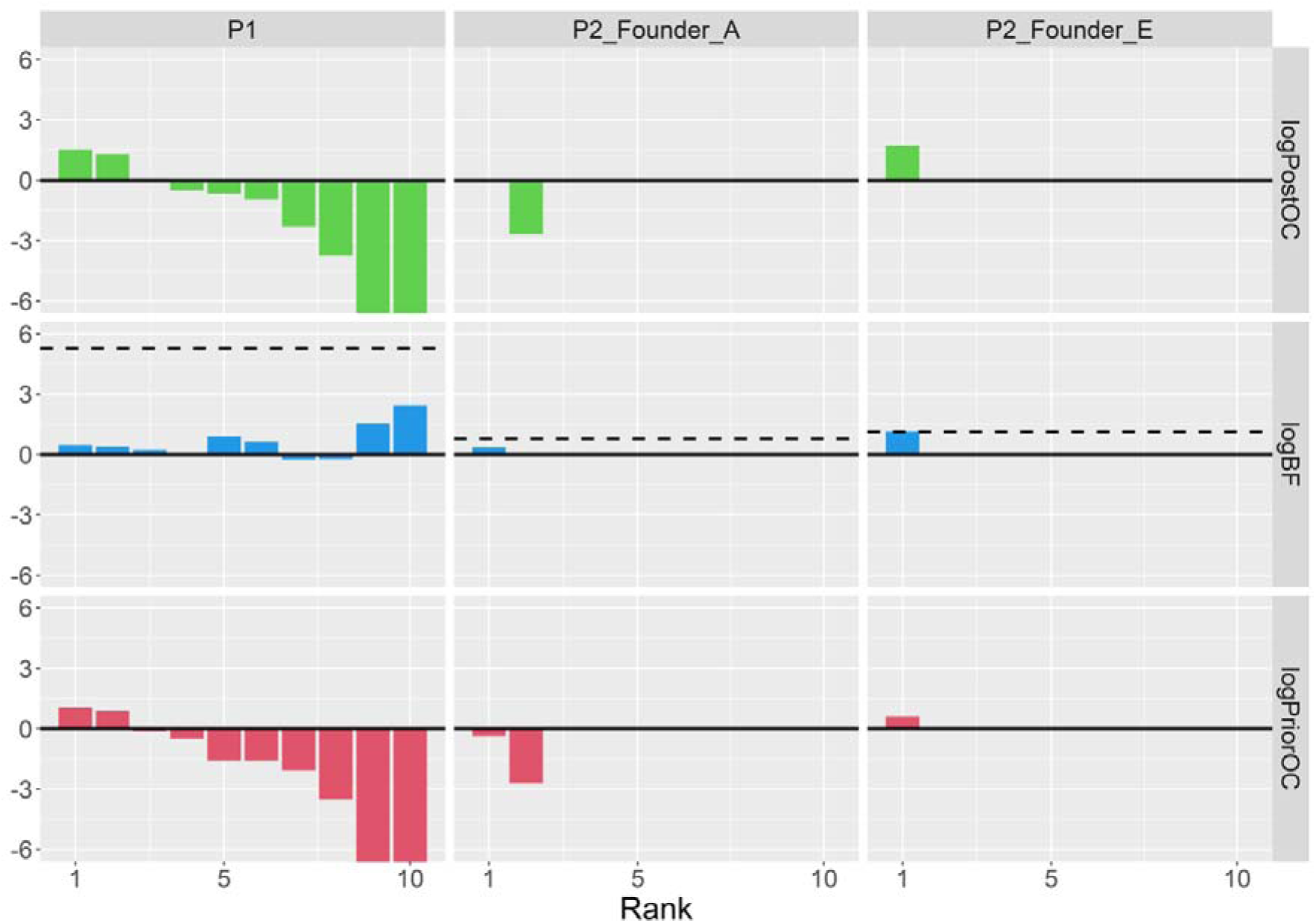
BICEP output metrics for the top 10 large, genic CNVs in the pedigrees/branches. The dashed line in the middle panel indicates the maximum logBF achievable in the pedigree/branch. For readability, the y-axis has been clipped at -6 due to common CNVs with very low logPriorOC and logPostOC values. Note that for the Founder A branch of P2, only two CNVs could be scored by BICEP, and in the Founder E branch, only one CNV could be scored.

#### 3.1.2 P2 pedigree

In the Founder A branch of P2, the top 8 ranked SNVs and indels were carried by three or fewer cases, making them poor candidates to explain OCD in this branch. The variant that ranked ninth, an ultra-rare, deleterious, missense variant in *MAP4*, had almost perfect co-segregation with OCD, being carried by five of the six cases for this branch (see Figure 3). In the Founder E branch, the third ranked variant achieved the maximum possible logBF in this pedigree. This was an ultra-rare missense variant in *MAPK8IP3* carried by all three cases (see Figure 4), although the evidence for deleteriousness was mixed (see Supplementary Table 3). In terms of known OCD risk genes, we observed one ultra-rare, deleterious missense variant in *BRWD1* that was carried by individual A. However, in the original publication, this gene was prioritised due to likely gene-disrupting variants, not due to deleterious missense variants, so this variant is unlikely to contribute to OCD risk in this individual.

**Figure 3:**
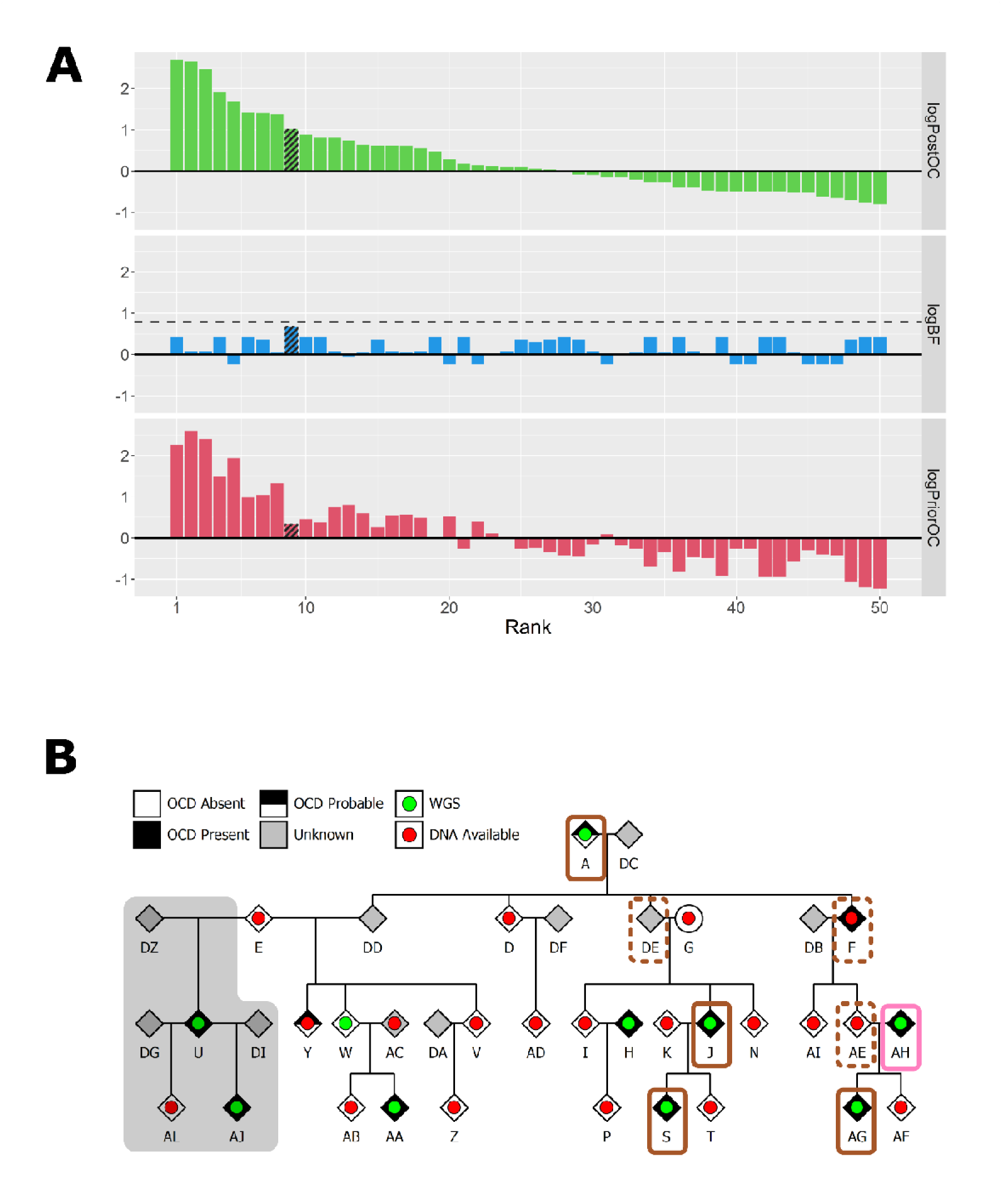
(**A**) BICEP output metrics for the top 50 ranked SNVs and indels in the Founder A branch of the P2 pedigree. The dashed line in the middle panels indicates the maximum logBF achievable. The shading represents the *MAP4* variant. (**B**) Pedigree diagram for the Founder A branch of P2. The coloured rectangles indicate carriers of the *MAP4* missense variant (brown), and the large deletion spanning 15q13.3 (pink). Solid borders represent observed variant carriers, and dashed borders represent obligate carriers. The grey background indicates the individuals from the other branch who were not included in these BICEP calculations.

**Figure 4:**
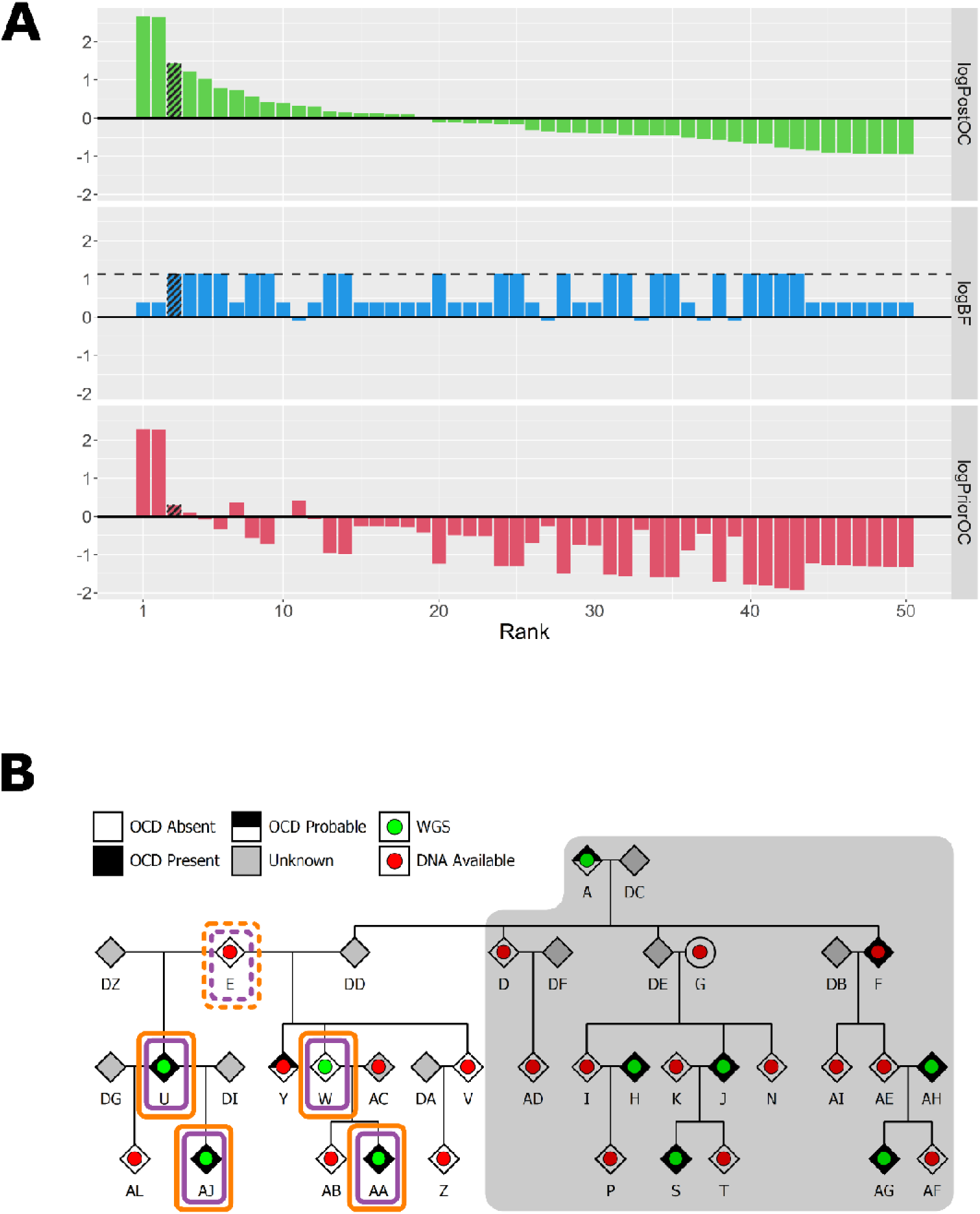
(**A**) BICEP output metrics for the top 50 ranked SNVs and indels in the Founder E branch of the P2 pedigree. The dashed line in the middle panels indicates the maximum logBF achievable. The shading represents the *MAPK8IP3* variant. (**B**) Pedigree diagram for the Founder E branch of P2. The coloured rectangles indicate carriers of the *MAPK8IP3* missense variant (purple), and the *DLGAP1* deletion (orange) Solid borders represent observed variant carriers, and dashed borders represent obligate carriers. The grey background indicates the individuals from the other branch who were not included in these BICEP calculations.

For CNVs, our calling and quality control resulted in 18 large, gene-disrupting CNVs (see Supplementary Table 4). In the Founder A branch, we did not observe any CNV with reasonable co-segregation patterns that also had positive logPriorOC scores (see Figure 2). However, in the Founder E branch, we observed a 947kbp deletion overlapping the first three exons in the untranslated region of *DLGAP1*. This CNV was called in all three cases and one control, achieving the maximum logBF for this branch. In all four carriers, the deletion was identified by at least three of the four CNV callers within PECAN, and a manual validation from the sequencing reads supported it as a true positive call (see Supplementary Figure 4). The deletion was absent from gnomAD and had a CADD-SV Phred score of 12.97, indicating that it is predicted to be in the top ∼5% most deleterious CNVs (Kleinert & Kircher, 2022).

Finally, we observed a large 23Mbp deletion overlapping the entire 15q13.3 critical region carried by individual AH (see Supplementary Figure 5). This individual has a diagnosis of OCD, but they are considered a married-in sample of the Founder A branch of the pedigree (see Figure 3). The deletion was not inherited by the child of AH, and this was the only known rare CNV observed in either branch of this pedigree.

We sought to identify protein-protein interactions between the four genes identified here and four genes that contained rare, deleterious variants segregating with TS in a previous study from our group (Ryan et al., 2022). A visualisation of this network from the STRING database is shown in Figure 5. *DLGAP1* and *MAPK8IP3* are connected to the previously reported cluster of *ERBB4* and *RAPGEF1*. No additional clustering between the input genes was observed, even when the size of the network was increased.

**Figure 5:**
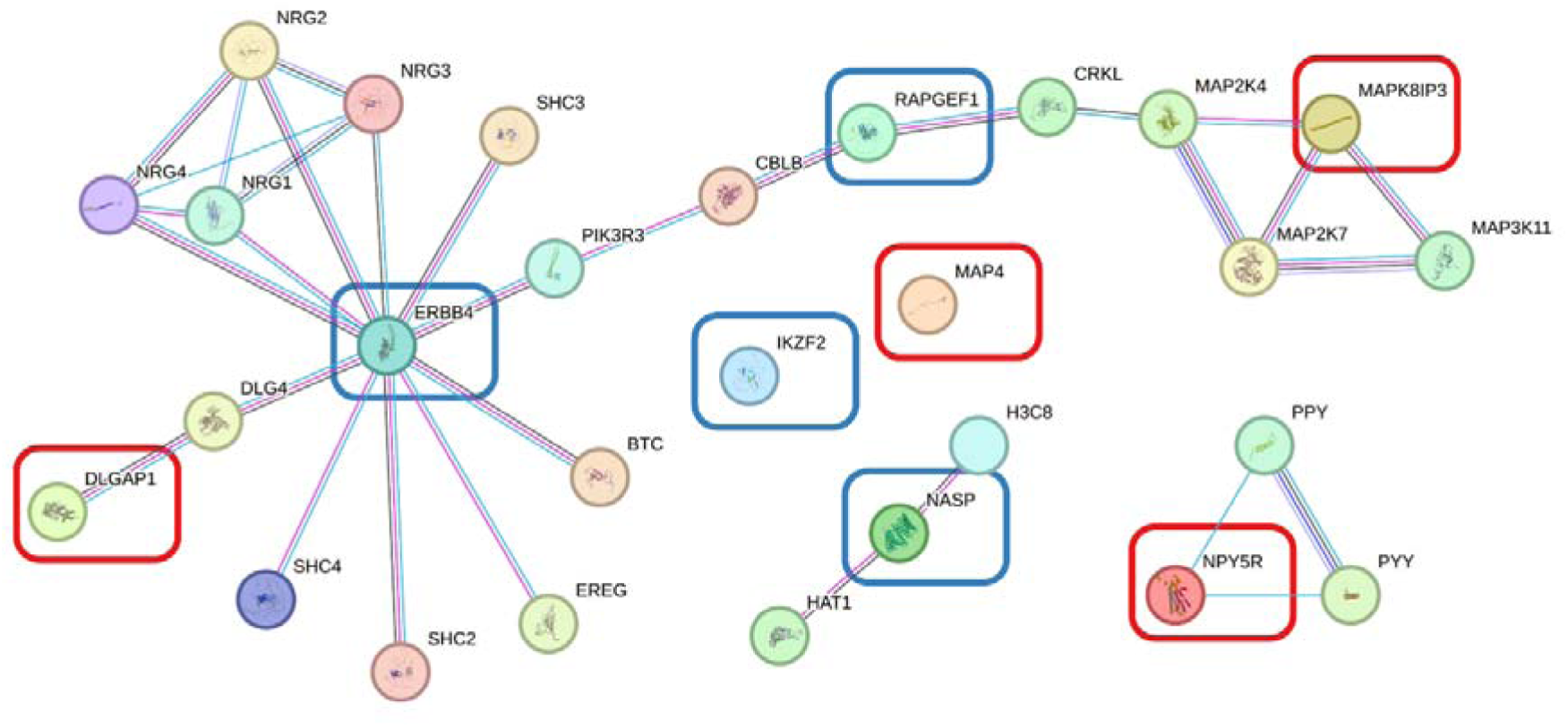
a protein-protein interaction network generated from STRING for the four genes identified in this study (red boxes) and the four genes identified from a previous family study of Tourette syndrome (blue boxes).

### 3.2 Common variants

We calculated PRS for three different traits for all pedigree members. We compared their PRS to a reference distribution generated from the EUR samples from the 1000 Genomes Project, as shown in Figure 6 below. We did not note any trends in the scores of the pedigree members, with a reasonable spread across the background distribution observed for all traits examined. In P1, the five non-carriers of the *NPY5R* variant did not have noticeably different PRS profiles compared to the carriers for any of the four selected traits.

**Figure 6:**
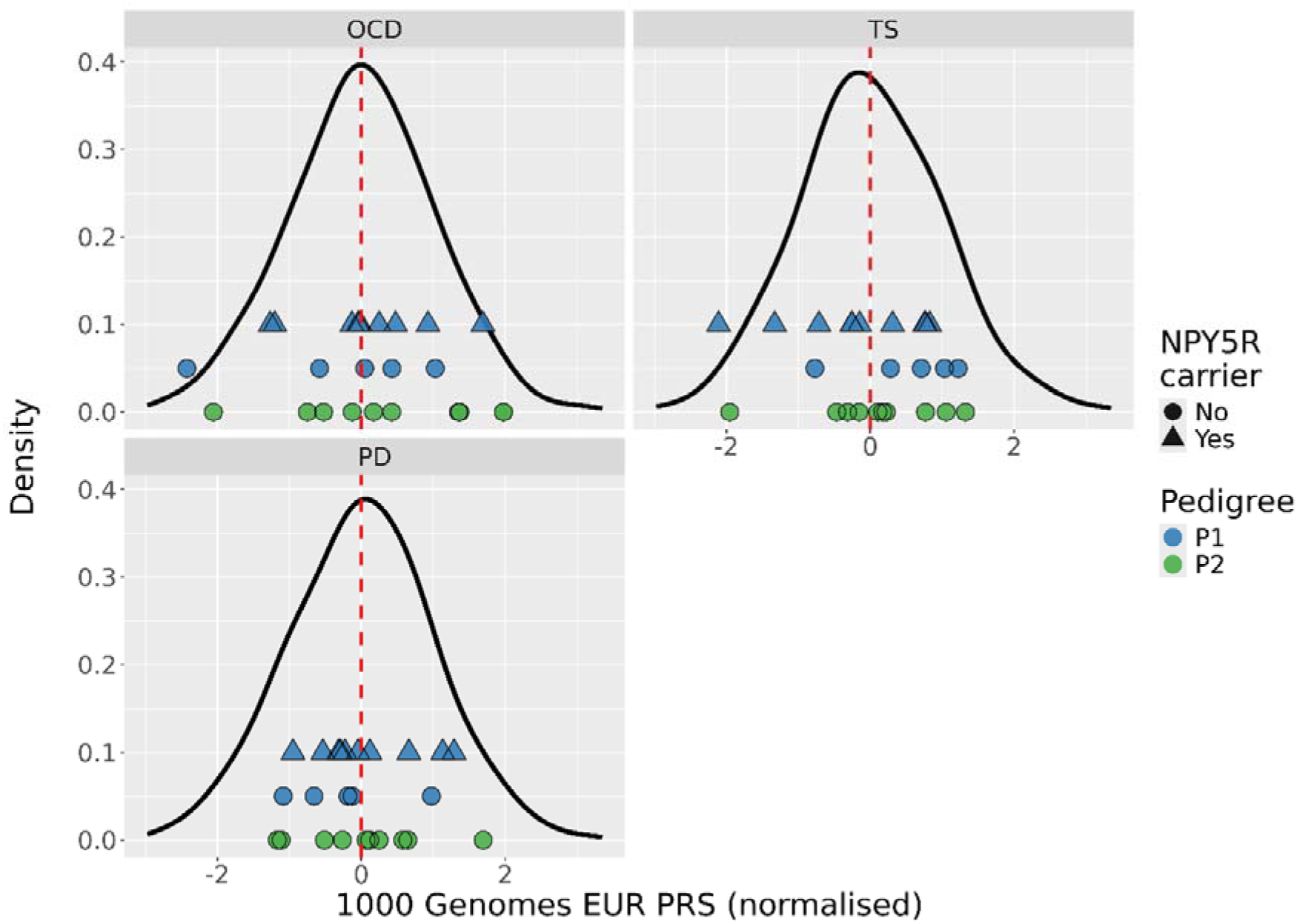
Density plots of the normalised PRS for the EUR reference samples (black line) for each of the three summary statistics. The blue and green points indicate PRS for the pedigree members, measured on the x-axis. The shape indicates carriers of the *NPY5R* missense variant. The dashed red line represents the mean PRS.

## 4 Discussion

In this study, we performed WGS on 25 samples from two multiplex OCD pedigrees, our aim being to identify rare, gene-disrupting variants that could explain the increased prevalence of OCD in these pedigrees. We examined a broad range of genomic variation making full use of the sequencing data. We focused on co-segregating variants, presented in order of their logPostOC scores from BICEP (see Supplementary Table 3). Here, we prioritise the genes that have shown previous association with OCD or its related neurobiology.

For pedigree P1, the most compelling evidence came from an ultra-rare highly deleterious missense variant in *NPY5R* carried by 10 out of the 15 pedigree cases. *NPY5R* is part of the Neuropeptide Y Receptor gene family, a brain expressed gene, hitherto implicated in feeding and stress regulation, which has been implicated in other anxiety phenotypes and depression (Eaton, Sallee, & Sah, 2007; Heilig, 2004; Roseboom et al., 2014). A role for common genetic variants at *NPY5R* have been proposed for panic disorder (Domschke et al., 2008; Otowa et al., 2009) and alcohol dependence (Wetherill et al., 2008). These findings have not been confirmed by larger GWAS studies of OCD or anxiety phenotypes, although significant loci in this gene have been reported in a GWAS of internet addiction (Haghighatfard et al., 2023). Several individuals in the pedigree reported anxiety-related disorders, including panic disorder (see Supplementary Table 1). As a secondary analysis, we re-ran BICEP on this pedigree with any anxiety disorder as the phenotype. However, the *NPY5R* variant did not show better co-segregation with anxiety than with OCD (see Supplementary Figure 6).

In pedigree P2, we noted that there were two main founders from which rare variants could likely be inherited, so we examined these two branches separately. In the Founder E branch of P2, we observed a large deletion of the untranslated region of *DLGAP1* that was also carried by all three cases in this branch. *DLGAP1* encodes for a post-synaptic density protein that is part of a family of scaffolding proteins that are involved in signalling to and from glutamate receptors. Several therapeutic agents involved in glutamatergic regulation have been proposal as experimental treatment for OCD (Grassi, Cecchelli, Vignozzi, & Pacini, 2020; Grassi, Scillitani, & Cecchelli, 2024). Variants in *DLGAP1* have been linked to both schizophrenia and autism in human studies and animal models (Coba et al., 2018; Rasmussen, Rasmussen, & Silahtaroglu, 2017). The gene has a pLI score of 1.00, which is consistent with previous work showing a strong enrichment of deletions disrupting loss-of-function intolerant genes in OCD (M. W. Halvorsen et al., 2025). *DLGAP1* has been implicated in OCD through rare CNVs identified in trios (Gazzellone et al., 2016) and from association studies (Arnold et al., 2018; Mattheisen et al., 2015; Stewart et al., 2013), although it was not reported in the most recent GWAS of OCD (Strom et al., 2025). However, we note that the majority of cases in this analysis were defined by self-report rather than clinically confirmed OCD cases. In a mouse model, *Dlgap1* knock out samples exhibited OCD-like behaviours at older ages (Minagawa et al., 2025).

Separate to the deletion, we identified a rare missense variant in *MAPK8IP3* that perfectly co-segregated with the sequenced cases. This variant was carried by the affected sample present in both branches. *MAPK8IP3* is a Mitogen-activated protein kinase that encodes for the JIP3 neuronal protein which is involved in axonal transport and neuronal development (Goldfarb et al., 2025). Recently, an epigenome-wide association analysis for OCD identified a significant methylation marker that was mapped to *MAPK8IP3* (Campos-Martin et al., 2023). More broadly, *de novo* variants in this gene have been reported in autism (Iossifov et al., 2014) and Smith-Magenis syndrome (Berger et al., 2017), and pathogenic variants in this gene are listed in the ClinVar database for complex neurodevelopmental disorders (Landrum et al., 2018).

Given the five cases in P1 not carrying any prioritised rare variant, we theorised that non-carriers in the cohort might have higher common variant burden, as might be expected under the liability threshold model (Q. Q. Huang et al., 2024). We generated PRS for OCD, TS, and PD, given the diagnoses present in the pedigree. However, we did not observe any trends in PRS between rare variant carriers and non-carriers (see Figure 6). In both families, there was a reasonable spread of pedigree members across the PRS distribution indicating that an increase in common variant burden is unlikely to be strongly influencing OCD risk in this cohort.

In this study, we have identified rare variants segregating with OCD in two multiplex pedigrees. The three genes highlighted here have shown previous association with OCD or related traits and are implicated in neuronal signalling and synaptic function. Two of the implicated genes show protein-protein interactions with other genes identified from a familial TS cohort, suggesting a possible molecular convergence between the two disorders on molecular mechanisms involving post-synaptic density (PSD) scaffolding and downstream neurodevelopmental processes relevant to synaptic plasticity and neuronal axonal transport. Some of these protein-disrupting variants are carried by a small number of cases and so will require replication in independent OCD cohorts. In addition, subsequent functional work may also validate their role as causal variants for OCD.

## Data Availability

All data produced in the present study are available upon reasonable request to the authors.

## 5 Acknowledgements

This work was supported by National Institutes of Health [5R01MH 124875 to A.C.].

## References

Altamura, C., Paluello, M. M., Mundo, E., Medda, S., & Mannu, P. (2001). Clinical and subclinical body dysmorphic disorder. Eur Arch Psychiatry Clin Neurosci, 251(3), 105–108. doi:10.1007/s004060170042

Arnold, P. D., Askland, K. D., Barlassina, C., Bellodi, L., Bienvenu, O. J., Black, D., . . . Zai, G. (2018). Revealing the complex genetic architecture of obsessive-compulsive disorder using meta-analysis. Mol. Psychiatry, 23(5), 1181–1188.

Auton, A., Brooks, L. D., Durbin, R. M., Garrison, E. P., Kang, H. M., Korbel, J. O., . . . Abecasis, G. R. (2015). A global reference for human genetic variation. Nature, 526(7571), 68–74.

Balachander, S., Meier, S., Matthiesen, M., Ali, F., Kannampuzha, A. J., Bhattacharya, M., . . . Viswanath, B. (2021). Are There Familial Patterns of Symptom Dimensions in Obsessive-Compulsive Disorder? Front Psychiatry, 12, 651196. doi:10.3389/fpsyt.2021.651196

Belyeu, J. R., Chowdhury, M., Brown, J., Pedersen, B. S., Cormier, M. J., Quinlan, A. R., & Layer, R. M. (2021). Samplot: a platform for structural variant visual validation and automated filtering. Genome Biol, 22(1), 161. doi:10.1186/s13059-021-02380-5

Berger, S. I., Ciccone, C., Simon, K. L., Malicdan, M. C., Vilboux, T., Billington, C., . . . Smith, A. C. M. (2017). Exome analysis of Smith-Magenis-like syndrome cohort identifies de novo likely pathogenic variants. Hum Genet, 136(4), 409–420. doi:10.1007/s00439-017-1767-x

Bienvenu, T., Lebrun, N., Clarke, J., Duriez, P., Gorwood, P., & Ramoz, N. (2019). Exome sequencing in a familial form of anorexia nervosa supports multigenic etiology. J Neural Transm (Vienna), 126(11), 1505–1511. doi:10.1007/s00702-019-02056-2

Blanco-Vieira, T., Radua, J., Marcelino, L., Bloch, M., Mataix-Cols, D., & do Rosário, M. C. (2023). The genetic epidemiology of obsessive-compulsive disorder: a systematic review and meta-analysis. Transl Psychiatry, 13(1), 230. doi:10.1038/s41398-023-02433-2

Bose, D., Fuchsberger, C., & Boehnke, M. (2025). Rare-variant association studies: When are aggregation tests more powerful than single-variant tests? Am J Hum Genet, 112(8), 1948–1961. doi:10.1016/j.ajhg.2025.07.002

Campos-Martin, R., Bey, K., Elsner, B., Reuter, B., Klawohn, J., Philipsen, A., . . . Ramirez, A. (2023). Epigenome-wide analysis identifies methylome profiles linked to obsessive-compulsive disorder, disease severity, and treatment response. Mol Psychiatry, 28(10), 4321–4330. doi:10.1038/s41380-023-02219-4

Cappi, C., Oliphant, M. E., Péter, Z., Zai, G., Conceição do Rosário, M., Sullivan, C. A. W., . . . Fernandez, T. V. (2020). De Novo Damaging DNA Coding Mutations Are Associated With Obsessive-Compulsive Disorder and Overlap With Tourette’s Disorder and Autism. Biol Psychiatry, 87(12), 1035–1044. doi:10.1016/j.biopsych.2019.09.029

Cavallini, M. C., Bertelli, S., Chiapparino, D., Riboldi, S., & Bellodi, L. (2000). Complex segregation analysis of obsessive-compulsive disorder in 141 families of eating disorder probands, with and without obsessive-compulsive disorder. Am J Med Genet, 96(3), 384–391. doi:10.1002/1096-8628(20000612)96:3<384::aid-ajmg28>3.0.co;2-p

Cavallini, M. C., Pasquale, L., Bellodi, L., & Smeraldi, E. (1999). Complex segregation analysis for obsessive compulsive disorder and related disorders. Am J Med Genet, 88(1), 38–43. doi:10.1002/(sici)1096-8628(19990205)88:1<38::aid-ajmg7>3.0.co;2-#

Cingolani, P., Patel, V. M., Coon, M., Nguyen, T., Land, S. J., Ruden, D. M., & Lu, X. (2012). Using Drosophila melanogaster as a Model for Genotoxic Chemical Mutational Studies with a New Program, SnpSift. Front Genet, 3, 35. doi:10.3389/fgene.2012.00035

Coba, M. P., Ramaker, M. J., Ho, E. V., Thompson, S. L., Komiyama, N. H., Grant, S. G. N., . . . Dulawa, S. C. (2018). Dlgap1 knockout mice exhibit alterations of the postsynaptic density and selective reductions in sociability. Sci Rep, 8(1), 2281. doi:10.1038/s41598-018-20610-y

Cui, H., Moore, J., Ashimi, S. S., Mason, B. L., Drawbridge, J. N., Han, S., . . . Lutter, M. (2013). Eating disorder predisposition is associated with ESRRA and HDAC4 mutations. J Clin Invest, 123(11), 4706–4713. doi:10.1172/jci71400

Domschke, K., Hohoff, C., Jacob, C., Maier, W., Fritze, J., Bandelow, B., . . . Deckert, J. (2008). Chromosome 4q31-34 panic disorder risk locus: association of neuropeptide Y Y5 receptor variants. Am J Med Genet B Neuropsychiatr Genet, 147b(4), 510-516. doi:10.1002/ajmg.b.30629

Eaton, K., Sallee, F. R., & Sah, R. (2007). Relevance of neuropeptide Y (NPY) in psychiatry. Curr Top Med Chem, 7(17), 1645–1659. doi:10.2174/156802607782341037

Ercan-Sencicek, A. G., Stillman, A. A., Ghosh, A. K., Bilguvar, K., O’Roak, B. J., Mason, C. E., . . . State, M. W. (2010). L-histidine decarboxylase and Tourette’s syndrome. N Engl J Med, 362(20), 1901–1908. doi:10.1056/NEJMoa0907006

First, M. B., Spitzer, R. L., Gibbon, M., & Williams, J. B. W. (2012). Structured Clinical Interview for DSM-IV® Axis I Disorders (SCID-I), Clinician Version, Administration Booklet: American Psychiatric Association Publishing.

Forstner, A. J., Awasthi, S., Wolf, C., Maron, E., Erhardt, A., Czamara, D., . . . Schumacher, J. (2021). Genome-wide association study of panic disorder reveals genetic overlap with neuroticism and depression. Mol Psychiatry, 26(8), 4179–4190. doi:10.1038/s41380-019-0590-2

Gazzellone, M. J., Zarrei, M., Burton, C. L., Walker, S., Uddin, M., Shaheen, S. M., . . . Scherer, S. W. (2016). Uncovering obsessive-compulsive disorder risk genes in a pediatric cohort by high-resolution analysis of copy number variation. J Neurodev Disord, 8, 36. doi:10.1186/s11689-016-9170-9

Ge, T., Chen, C. Y., Ni, Y., Feng, Y. A., & Smoller, J. W. (2019). Polygenic prediction via Bayesian regression and continuous shrinkage priors. Nat Commun, 10(1), 1776. doi:10.1038/s41467-019-09718-5

Glahn, D. C., Nimgaonkar, V. L., Raventos, H., Contreras, J., McIntosh, A. M., Thomson, P. A., . . . Blangero, J. (2019). Rediscovering the value of families for psychiatric genetics research. Mol. Psychiatry, 24(4), 523–535.

Goldfarb, T., Kodali, V. K., Pujar, S., Brover, V., Robbertse, B., Farrell, C. M., . . . Murphy, T. D. (2025). NCBI RefSeq: reference sequence standards through 25 years of curation and annotation. Nucleic Acids Res, 53(D1), D243–d257. doi:10.1093/nar/gkae1038

Goodman, W. K., Price, L. H., Rasmussen, S. A., Mazure, C., Fleischmann, R. L., Hill, C. L., . . . Charney, D. S. (1989). The Yale-Brown Obsessive Compulsive Scale. I. Development, use, and reliability. Arch Gen Psychiatry, 46(11), 1006–1011. doi:10.1001/archpsyc.1989.01810110048007

Grassi, G., Cecchelli, C., Vignozzi, L., & Pacini, S. (2020). Investigational and Experimental Drugs to Treat Obsessive-Compulsive Disorder. J Exp Pharmacol, 12, 695–706. doi:10.2147/jep.S255375

Grassi, G., Scillitani, E., & Cecchelli, C. (2024). New horizons for obsessive-compulsive disorder drug discovery: is targeting glutamate receptors the answer? Expert Opin Drug Discov, 19(10), 1235–1245. doi:10.1080/17460441.2024.2387127

Grotzinger, A. D., Werme, J., Peyrot, W. J., Frei, O., de Leeuw, C., Bicks, L. K., . . . Smoller, J. W. (2026). Mapping the genetic landscape across 14 psychiatric disorders. Nature, 649(8096), 406–415. doi:10.1038/s41586-025-09820-3

Haeussler, M., Zweig, A. S., Tyner, C., Speir, M. L., Rosenbloom, K. R., Raney, B. J., . . . Kent, W. J. (2019). The UCSC Genome Browser database: 2019 update. Nucleic Acids Res, 47(D1), D853–d858. doi:10.1093/nar/gky1095

Haghighatfard, A., Ghaderi, A. H., Mostajabi, P., Kashfi, S. S., Mohabati Somehsarayee, H., Shahrani, M., . . . Moghadam, E. R. (2023). The first genome-wide association study of internet addiction; Revealed substantial shared risk factors with neurodevelopmental psychiatric disorders. Res Dev Disabil, 133, 104393. doi:10.1016/j.ridd.2022.104393

Halvorsen, M., Samuels, J., Wang, Y., Greenberg, B. D., Fyer, A. J., McCracken, J. T., . . . Goldstein, D. B. (2021). Exome sequencing in obsessive-compulsive disorder reveals a burden of rare damaging coding variants. Nat Neurosci, 24(8), 1071–1076. doi:10.1038/s41593-021-00876-8

Halvorsen, M. W., de Schipper, E., Bäckman, J., Strom, N. I., Hagen, K., Lindblad-Toh, K., . . . Crowley, J. J. (2025). A burden of rare copy number variants in obsessive-compulsive disorder. Mol Psychiatry, 30(4), 1510–1517. doi:10.1038/s41380-024-02763-7

Heilig, M. (2004). The NPY system in stress, anxiety and depression. Neuropeptides, 38(4), 213–224. doi:10.1016/j.npep.2004.05.002

Huang, A. Y., Yu, D., Davis, L. K., Sul, J. H., Tsetsos, F., Ramensky, V., . . . Coppola, G. (2017). Rare Copy Number Variants in NRXN1 and CNTN6 Increase Risk for Tourette Syndrome. Neuron, 94(6), 1101–1111.e1107. doi:10.1016/j.neuron.2017.06.010

Huang, M. H., Cheng, C. M., Tsai, S. J., Bai, Y. M., Li, C. T., Lin, W. C., Chen, M. H. (2021). Familial coaggregation of major psychiatric disorders among first-degree relatives of patients with obsessive-compulsive disorder: a nationwide study. Psychol Med, 51(4), 680–687. doi:10.1017/s0033291719003696

Huang, Q. Q., Wigdor, E. M., Malawsky, D. S., Campbell, P., Samocha, K. E., Chundru, V. K., . . . Martin, H. C. (2024). Examining the role of common variants in rare neurodevelopmental conditions. Nature, 636(8042), 404–411. doi:10.1038/s41586-024-08217-y

Iossifov, I., O’Roak, B. J., Sanders, S. J., Ronemus, M., Krumm, N., Levy, D., . . . Wigler, M. (2014). The contribution of de novo coding mutations to autism spectrum disorder. Nature, 515(7526), 216–221. doi:10.1038/nature13908

Karczewski, K. J., Francioli, L. C., Tiao, G., Cummings, B. B., Alföldi, J., Wang, Q., . . . MacArthur, D. G. (2020). The mutational constraint spectrum quantified from variation in 141,456 humans. Nature, 581(7809), 434–443. doi:10.1038/s41586-020-2308-7

Kaufman, J., Birmaher, B., Brent, D., Rao, U., Flynn, C., Moreci, P., . . . Ryan, N. (1997). Schedule for Affective Disorders and Schizophrenia for School-Age Children-Present and Lifetime Version (K-SADS-PL): initial reliability and validity data. J Am Acad Child Adolesc Psychiatry, 36(7), 980–988. doi:10.1097/00004583-199707000-00021

Kendall, K. M., Bracher-Smith, M., Fitzpatrick, H., Lynham, A., Rees, E., Escott-Price, V., . . . Kirov, G. (2019). Cognitive performance and functional outcomes of carriers of pathogenic copy number variants: analysis of the UK Biobank. Br J Psychiatry, 214(5), 297–304. doi:10.1192/bjp.2018.301

Kleinert, P., & Kircher, M. (2022). A framework to score the effects of structural variants in health and disease. Genome Res, 32(4), 766–777. doi:10.1101/gr.275995.121

Landrum, M. J., Lee, J. M., Benson, M., Brown, G. R., Chao, C., Chitipiralla, S., . . . Maglott, D. R. (2018). ClinVar: improving access to variant interpretations and supporting evidence. Nucleic Acids Res, 46(D1), D1062–d1067. doi:10.1093/nar/gkx1153

Lee, S., Abecasis, G. R., Boehnke, M., & Lin, X. (2014). Rare-variant association analysis: study designs and statistical tests. Am J Hum Genet, 95(1), 5–23. doi:10.1016/j.ajhg.2014.06.009

Manichaikul, A., Mychaleckyj, J. C., Rich, S. S., Daly, K., Sale, M., & Chen, W. M. (2010). Robust relationship inference in genome-wide association studies. Bioinformatics, 26(22), 2867–2873. doi:10.1093/bioinformatics/btq559

Marshall, C. R., Howrigan, D. P., Merico, D., Thiruvahindrapuram, B., Wu, W., Greer, D. S., . . . Sebat, J. (2017). Contribution of copy number variants to schizophrenia from a genome-wide study of 41,321 subjects. Nat Genet, 49(1), 27–35. doi:10.1038/ng.3725

Mathews, C. A., Badner, J. A., Andresen, J. M., Sheppard, B., Himle, J. A., Grant, J. E., . . . Hanna, G. L. (2012). Genome-wide linkage analysis of obsessive-compulsive disorder implicates chromosome 1p36. Biol Psychiatry, 72(8), 629–636. doi:10.1016/j.biopsych.2012.03.037

Mathews, C. A., & Grados, M. A. (2011). Familiality of Tourette syndrome, obsessive-compulsive disorder, and attention-deficit/hyperactivity disorder: heritability analysis in a large sib-pair sample. J Am Acad Child Adolesc Psychiatry, 50(1), 46–54. doi:10.1016/j.jaac.2010.10.004

Mathews, C. A., Nievergelt, C. M., Azzam, A., Garrido, H., Chavira, D. A., Wessel, J., . . . Schork, N. J. (2007). Heritability and clinical features of multigenerational families with obsessive-compulsive disorder and hoarding. Am J Med Genet B Neuropsychiatr Genet, 144b(2), 174-182. doi:10.1002/ajmg.b.30370

Mattheisen, M., Samuels, J. F., Wang, Y., Greenberg, B. D., Fyer, A. J., McCracken, J. T., . . . Nestadt, G. (2015). Genome-wide association study in obsessive-compulsive disorder: results from the OCGAS. Mol Psychiatry, 20(3), 337–344. doi:10.1038/mp.2014.43

McGrath, L. M., Yu, D., Marshall, C., Davis, L. K., Thiruvahindrapuram, B., Li, B., . . . Scharf, J. M. (2014). Copy number variation in obsessive-compulsive disorder and tourette syndrome: a cross-disorder study. J Am Acad Child Adolesc Psychiatry, 53(8), 910–919. doi:10.1016/j.jaac.2014.04.022

McLaren, W., Gil, L., Hunt, S. E., Riat, H. S., Ritchie, G. R., Thormann, A., . . . Cunningham, F. (2016). The Ensembl Variant Effect Predictor. Genome Biol, 17(1), 122. doi:10.1186/s13059-016-0974-4

Minagawa, K., Hayakawa, T., Akimoto, H., Nagashima, T., Takahashi, Y., & Asai, S. (2025). Late development of OCD-like phenotypes in Dlgap1 knockout mice. Psychopharmacology (Berl), 242(1), 215–231. doi:10.1007/s00213-024-06668-9

Morales, J., Pujar, S., Loveland, J. E., Astashyn, A., Bennett, R., Berry, A., . . . Murphy, T. D. (2022). A joint NCBI and EMBL-EBI transcript set for clinical genomics and research. Nature, 604(7905), 310–315. doi:10.1038/s41586-022-04558-8

Nestadt, G., Lan, T., Samuels, J., Riddle, M., Bienvenu, O. J., 3rd, Liang, K. Y., . . . Shugart, Y. Y. (2000). Complex segregation analysis provides compelling evidence for a major gene underlying obsessive-compulsive disorder and for heterogeneity by sex. Am J Hum Genet, 67(6), 1611-1616. doi:10.1086/316898

Nestadt, G., Samuels, J., Riddle, M., Bienvenu, O. J., 3rd, Liang, K. Y., LaBuda, M., . . . Hoehn-Saric, R. (2000). A family study of obsessive-compulsive disorder. Arch Gen Psychiatry, 57(4), 358-363. doi:10.1001/archpsyc.57.4.358

Nurnberger, J. I., Jr., Blehar, M. C., Kaufmann, C. A., York-Cooler, C., Simpson, S. G., Harkavy-Friedman, J., . . . Reich, T. (1994). Diagnostic interview for genetic studies. Rationale, unique features, and training. NIMH Genetics Initiative. Arch Gen Psychiatry, 51(11), 849–859; discussion 863-844. doi:10.1001/archpsyc.1994.03950110009002

Ormond, C., Ryan, N. M., Byerley, W., Heron, E. A., & Corvin, A. (2024). Investigating copy number variants in schizophrenia pedigrees using a new consensus pipeline called PECAN. Sci Rep, 14(1), 17518. doi:10.1038/s41598-024-66021-0

Ormond, C., Ryan, N. M., Cap, M., Byerley, W., Corvin, A., & Heron, E. A. (2024). BICEP: Bayesian inference for rare genomic variant causality evaluation in pedigrees. Brief Bioinform, 26(1). doi:10.1093/bib/bbae624

Ormond, C., Ryan, N. M., Corvin, A., & Heron, E. A. (2021). Converting single nucleotide variants between genome builds: from cautionary tale to solution. Brief Bioinform, 22(5). doi:10.1093/bib/bbab069

Ormond, C., Ryan, N. M., Corvin, A., & Heron, E. A. (2026). BICEP: an extension to indels and copy number variants for rare variant prioritisation in pedigree analysis. bioRxiv, 2026.2003.2009.710467. doi:10.64898/2026.03.09.710467

Ormond, C., Ryan, N. M., Heron, E. A., Gill, M., Byerley, W., & Corvin, A. (2023). Ultra-rare missense variants implicated in Utah pedigrees multiply affected with schizophrenia. Biological Psychiatry: Global Open Science. doi:10.1016/j.bpsgos.2023.02.002

Orvaschel, H., & Puig-Antich, J. (1987). Schedule for affective disorders and schizophrenia for school-age children: Epidemiologic version. Fort Lauderdale, FL: Nova University.

Otowa, T., Yoshida, E., Sugaya, N., Yasuda, S., Nishimura, Y., Inoue, K., . . . Okazaki, Y. (2009). Genome-wide association study of panic disorder in the Japanese population. J Hum Genet, 54(2), 122–126. doi:10.1038/jhg.2008.17

Pauls, D., & Hurst, C. (1987). Schedule for Tourette’s syndrome and other behavioral syndromes. New Haven, CT: Yale University Child Study Center.

Pedersen, B. S., & Quinlan, A. R. (2017). Who’s Who? Detecting and Resolving Sample Anomalies in Human DNA Sequencing Studies with Peddy. Am J Hum Genet, 100(3), 406–413. doi:10.1016/j.ajhg.2017.01.017

Privé, F., Aschard, H., Ziyatdinov, A., & Blum, M. G. B. (2018). Efficient analysis of large-scale genome-wide data with two R packages: bigstatsr and bigsnpr. Bioinformatics, 34(16), 2781–2787. doi:10.1093/bioinformatics/bty185

Purcell, S., Neale, B., Todd-Brown, K., Thomas, L., Ferreira, M. A., Bender, D., . . . Sham, P. C. (2007). PLINK: a tool set for whole-genome association and population-based linkage analyses. Am J Hum Genet, 81(3), 559–575. doi:10.1086/519795

Rasmussen, A. H., Rasmussen, H. B., & Silahtaroglu, A. (2017). The DLGAP family: neuronal expression, function and role in brain disorders. Mol Brain, 10(1), 43. doi:10.1186/s13041-017-0324-9

Rees, E., & Kirov, G. (2021). Copy number variation and neuropsychiatric illness. Curr Opin Genet Dev, 68, 57–63. doi:10.1016/j.gde.2021.02.014

Rees, E., Walters, J. T., Chambert, K. D., O’Dushlaine, C., Szatkiewicz, J., Richards, A. L., . . . Kirov, G. (2014). CNV analysis in a large schizophrenia sample implicates deletions at 16p12.1 and SLC1A1 and duplications at 1p36.33 and CGNL1. Hum Mol Genet, 23(6), 1669–1676. doi:10.1093/hmg/ddt540

Rees, E., Walters, J. T., Georgieva, L., Isles, A. R., Chambert, K. D., Richards, A. L., . . . Kirov, G. (2014). Analysis of copy number variations at 15 schizophrenia-associated loci. Br J Psychiatry, 204(2), 108–114. doi:10.1192/bjp.bp.113.131052

Roseboom, P. H., Nanda, S. A., Fox, A. S., Oler, J. A., Shackman, A. J., Shelton, S. E., . . . Kalin, N. H. (2014). Neuropeptide Y receptor gene expression in the primate amygdala predicts anxious temperament and brain metabolism. Biol Psychiatry, 76(11), 850–857. doi:10.1016/j.biopsych.2013.11.012

Ross, J., Badner, J., Garrido, H., Sheppard, B., Chavira, D. A., Grados, M., . . . Mathews, C. A. (2011). Genomewide linkage analysis in Costa Rican families implicates chromosome 15q14 as a candidate region for OCD. Hum Genet, 130(6), 795–805. doi:10.1007/s00439-011-1033-6

Ruscio, A. M., Stein, D. J., Chiu, W. T., & Kessler, R. C. (2010). The epidemiology of obsessive-compulsive disorder in the National Comorbidity Survey Replication. Mol Psychiatry, 15(1), 53–63. doi:10.1038/mp.2008.94

Ryan, N. M., Ormond, C., Chang, Y. C., Contreras, J., Raventos, H., Gill, M., . . . Corvin, A. (2022). Identity-by-descent analysis of a large Tourette’s syndrome pedigree from Costa Rica implicates genes involved in neuronal development and signal transduction. Mol Psychiatry. doi:10.1038/s41380-022-01771-9

Samuels, J. F., Bienvenu, O. J., 3rd, Pinto, A., Fyer, A. J., McCracken, J. T., Rauch, S. L., . . . Nestadt, G. (2007). Hoarding in obsessive-compulsive disorder: results from the OCD Collaborative Genetics Study. Behav Res Ther, 45(4), 673-686. doi:10.1016/j.brat.2006.05.008

Sanders, S. J. (2015). First glimpses of the neurobiology of autism spectrum disorder. Curr. Opin. Genet. Dev., 33, 80–92.

Scahill, L., Riddle, M. A., McSwiggin-Hardin, M., Ort, S. I., King, R. A., Goodman, W. K., . . . Leckman, J. F. (1997). Children’s Yale-Brown Obsessive Compulsive Scale: reliability and validity. J Am Acad Child Adolesc Psychiatry, 36(6), 844–852. doi:10.1097/00004583-199706000-00023

Shanta, O., Klein, M., Sacks, M., MacDonald, J. R., Maihofer, A., Ahangari, M., . . . Sebat, J. (2025). A cross-disorder analysis of CNVs finds novel loci and dose-dependent relationships of genes to psychiatric traits. medRxiv. doi:10.1101/2025.07.11.25331310

Stewart, S. E., Yu, D., Scharf, J. M., Neale, B. M., Fagerness, J. A., Mathews, C. A., . . . Pauls, D. L. (2013). Genome-wide association study of obsessive-compulsive disorder. Mol Psychiatry, 18(7), 788–798. doi:10.1038/mp.2012.85

Strom, N. I., Gerring, Z. F., Galimberti, M., Yu, D., Halvorsen, M. W., Abdellaoui, A., . . . Mattheisen, M. (2025). Genome-wide analyses identify 30 loci associated with obsessive-compulsive disorder. Nat Genet, 57(6), 1389–1401. doi:10.1038/s41588-025-02189-z

Sun, N., Nasello, C., Deng, L., Wang, N., Zhang, Y., Xu, Z., . . . Tischfield, J. A. (2018). The PNKD gene is associated with Tourette Disorder or Tic disorder in a multiplex family. Mol Psychiatry, 23(6), 1487–1495. doi:10.1038/mp.2017.179

Sundaram, S. K., Huq, A. M., Sun, Z., Yu, W., Bennett, L., Wilson, B. J., . . . Chugani, H. T. (2011). Exome sequencing of a pedigree with Tourette syndrome or chronic tic disorder. Ann Neurol, 69(5), 901–904. doi:10.1002/ana.22398

Szklarczyk, D., Kirsch, R., Koutrouli, M., Nastou, K., Mehryary, F., Hachilif, R., . . . von Mering, C. (2023). The STRING database in 2023: protein-protein association networks and functional enrichment analyses for any sequenced genome of interest. Nucleic Acids Res, 51(D1), D638–d646. doi:10.1093/nar/gkac1000

Wang, B., Tran, M. N., Wang, S., Liu, Y., Olfson, E., Wang, G., . . . State, M. W. (2025). Rare coding mutations identify 36 large-effect risk genes in obsessive-compulsive disorder and chronic tic disorders. medRxiv. doi:10.1101/2025.10.10.25337672

Wetherill, L., Schuckit, M. A., Hesselbrock, V., Xuei, X., Liang, T., Dick, D. M., . . . Foroud, T. (2008). Neuropeptide Y receptor genes are associated with alcohol dependence, alcohol withdrawal phenotypes, and cocaine dependence. Alcohol Clin Exp Res, 32(12), 2031–2040. doi:10.1111/j.1530-0277.2008.00790.x

Willour, V. L., Yao Shugart, Y., Samuels, J., Grados, M., Cullen, B., Bienvenu, O. J., 3rd, . . . Nestadt, G. (2004). Replication study supports evidence for linkage to 9p24 in obsessive-compulsive disorder. Am J Hum Genet, 75(3), 508-513. doi:10.1086/423899

Yu, D., Sul, J. H., Tsetsos, F., Nawaz, M. S., Huang, A. Y., Zelaya, I., . . . Scharf, J. M. (2019). Interrogating the Genetic Determinants of Tourette’s Syndrome and Other Tic Disorders Through Genome-Wide Association Studies. Am J Psychiatry, 176(3), 217–227. doi:10.1176/appi.ajp.2018.18070857

Zhou, W., Nielsen, J. B., Fritsche, L. G., Dey, R., Gabrielsen, M. E., Wolford, B. N., . . . Lee, S. (2018). Efficiently controlling for case-control imbalance and sample relatedness in large-scale genetic association studies. Nat Genet, 50(9), 1335–1341. doi:10.1038/s41588-018-0184-y

