## Supplementary materials for "Rare protein-disrupting variants in *NPY5R, DLGAP1* and *MAPK8IP3* segregate with OCD in two multiplex pedigrees"

**Affiliations**

### 1 Supplementary Methods

#### 1.1 Pedigree consistency

For the P2 pedigree, the WGS data indicated inconsistent relatedness compared to the original pedigree diagram. SNP-array data were also available for all samples with DNA in this cohort. The array data was converted from genome build GRCh36 to build GRCh38 using liftOver (Haeussler et al., 2019). We first identified conversion-unstable positions (Cathal Ormond, Ryan, Corvin, & Heron, 2021) from genome build GRCh36 to GRCh38 using the chain files from Ensembl (see Supplementary Methods). This process generated no novel unstable positions, just positions which could not be converted from build GRCh36. Following conversion, SNPs were renamed and compared to variants in the WGS data. Variants shared by both datasets were extracted, and the two datasets were merged. Using plink (Purcell et al., 2007), we confirmed that no sample had excess heterozygosity (F > 0.2) or missingness (F_MISS > 0.02). Next, we removed SNPs with excessive missingness (F_MISS > 0.02) or Hardy-Weinburg equilibrium violations ($p<{10}^{-6}$ for cases and $p<{10}^{-10}$ for controls). Finally, we generated a list of LD-pruned SNPs (--indep-pairwise 100 50 0.2) that were common (minor allele frequency > 0.05). Pairwise relatedness coefficients were generated on this subset of SNPs using plink (Purcell et al., 2007).

These scores showed perfect concordance between the WGS and array data (indicating there were no sample ID mismatches) and showed the same relatedness issues as before. The full pedigree genomic data indicated that sample U was a half sibling instead of a full sibling to samples Y, W, and V (see Supplementary Figure 2). When this change was made to the pedigree diagram, all observed pairwise relatedness coefficients matched the expected pedigree relatedness.

#### 1.2 Prior model benchmarking

For missense variants, we explored using PolyPhen2 as the sole deleteriousness score, instead of the five default deleteriousness scores (which includes PolyPhen2). We evaluated the predictive accuracy of the two prior models for missense variants by splitting the ClinVar regression data into training (80%) and testing (20%) subsets. As described in the original BICEP publication (C. Ormond et al., 2024), we calculated five performance metrics on both datasets: sensitivity, specificity, the positive predictive value (PPV), the negative predictive value (NPV) and the Matthews correlation coefficient (MCC). We generated central 95% confidence intervals by bootstrapping.

#### 1.3 Web resources

- Ensembl chain files:

<https://ftp.ensembl.org/pub/assembly_mapping/homo_sapiens/>

- 1000 Genomes high coverage WGS data:

<https://ftp.1000genomes.ebi.ac.uk/vol1/ftp/data_collections/1000G_2504_high_coverage/working/20220422_3202_phased_SNV_INDEL_SV/>

- Psychiatric Genomics Consortium summary statistics:

<https://pgc.unc.edu/for-researchers/download-results/>

### 2 Supplementary Figures

(**A**)
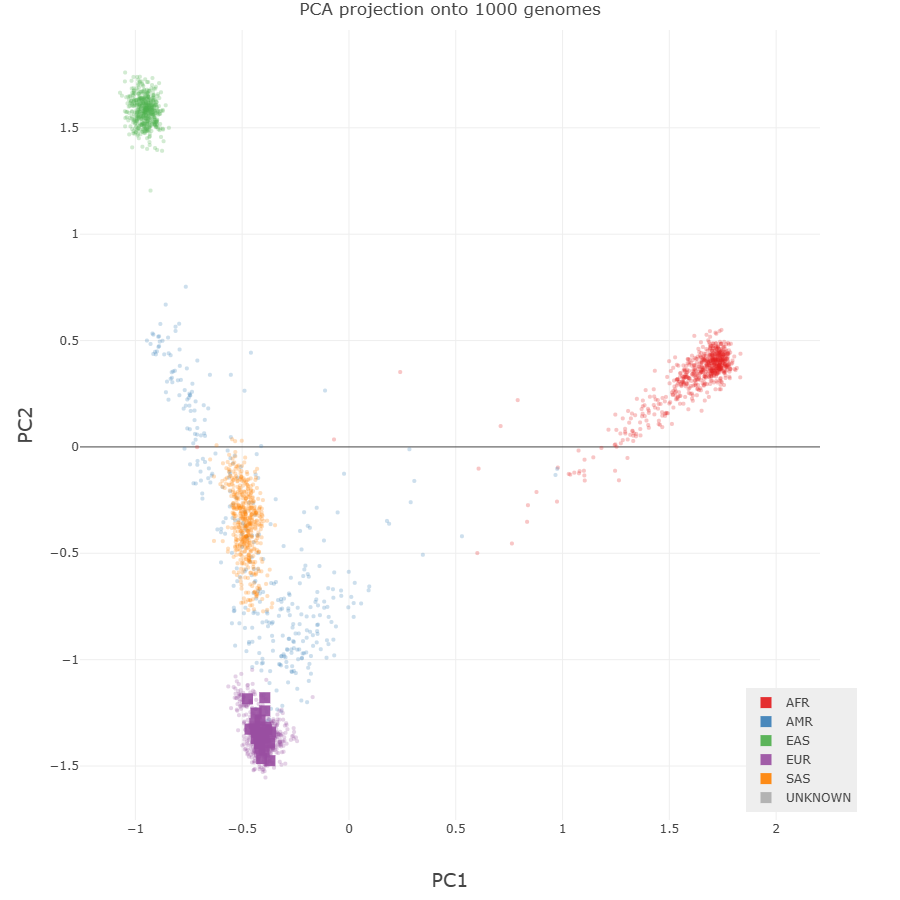

(**B**)

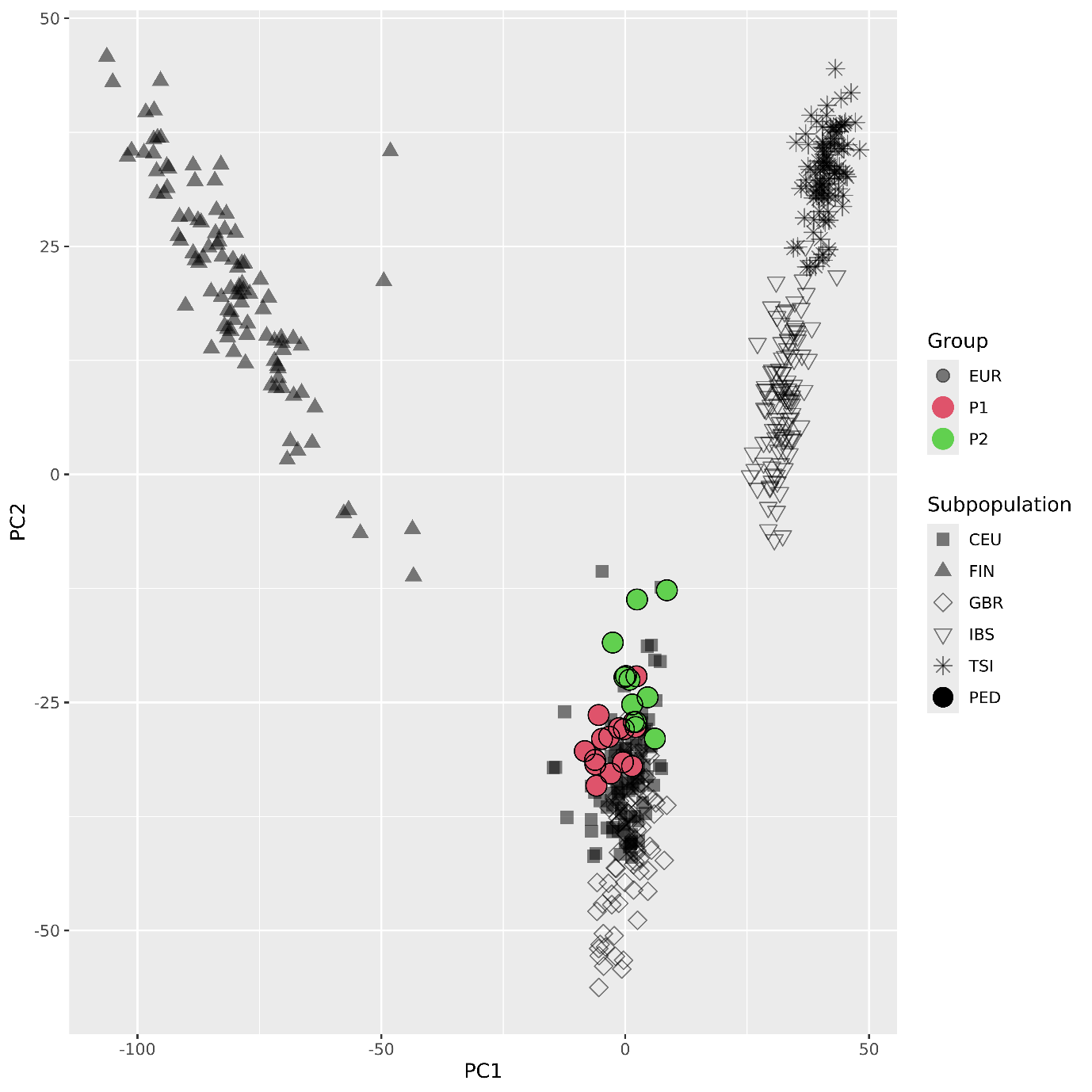

Supplementary Figure 1: (**A**) PCA output from peddy (Pedersen & Quinlan, 2017) with the entire 1000 Genomes Project samples, coloured by genomic ancestry. Dots represent the reference samples, and squares represent pedigree members. (**B**) PCA output from “bigsnpr” using the EUR subset of the 1000 Genomes Project samples. The shapes represent the reference sub-populations within the EUR cluster, and the colours represent the pedigree members (PED).

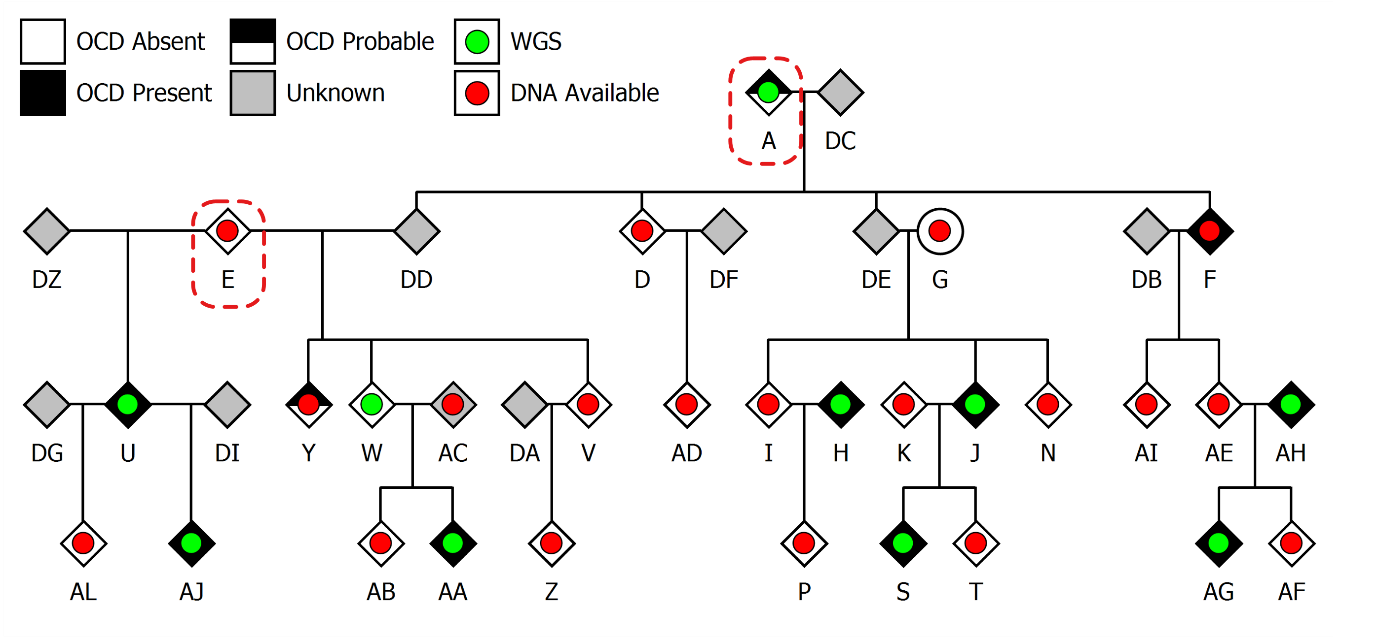

Supplementary Figure 2: Pedigree P2. The two unrelated founders (individuals A and E) are marked with the red dashed boxes. The legend indicates OCD status, DNA availability, and the samples selected for sequencing.

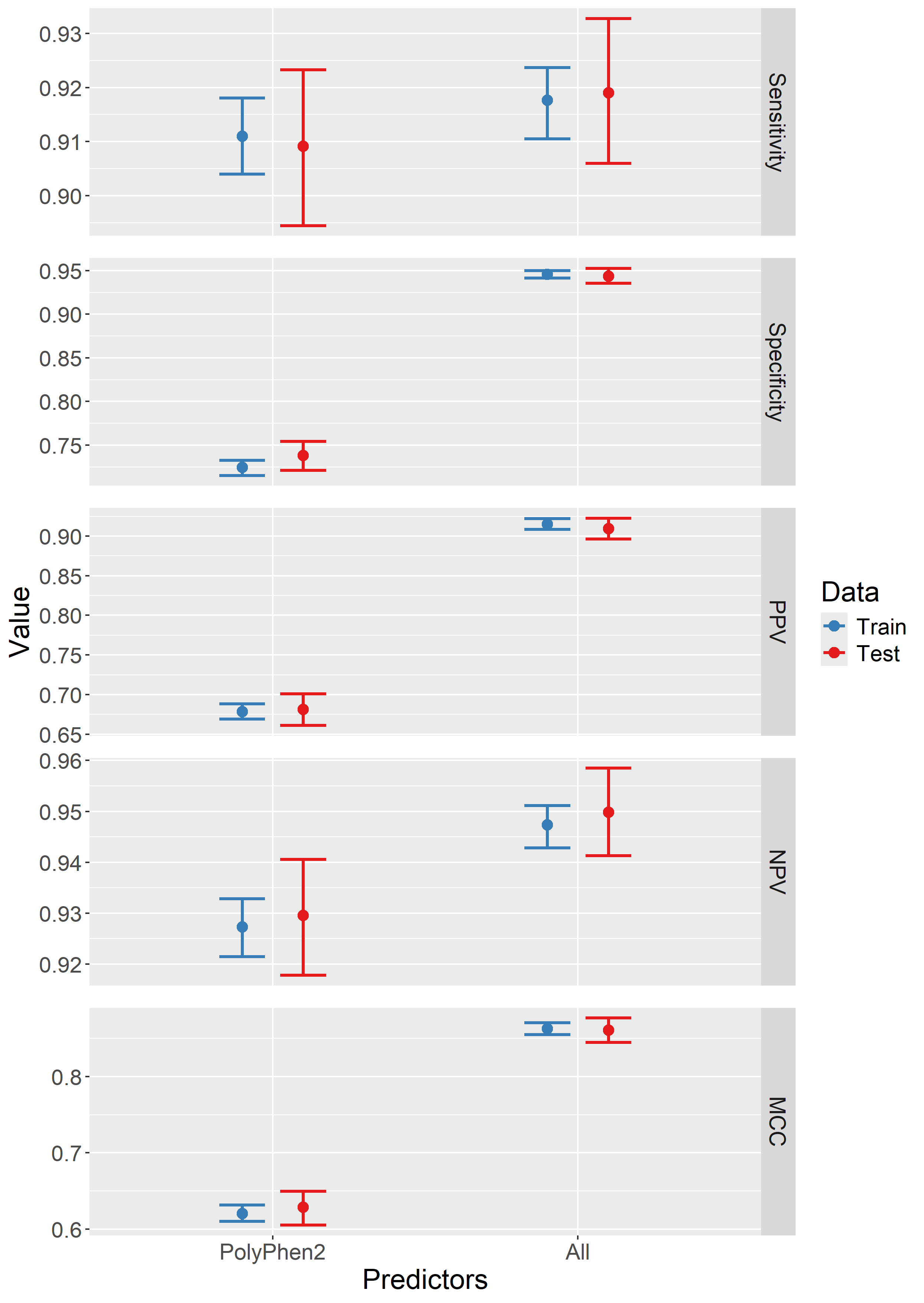

Supplementary Figure 3: BICEP benchmark metrics for the two different prior models for missense variants (just PolyPhen2, and all five deleteriousness predictors in the original model). Metrics for the training and test datasets are supplied with central 95% confidence intervals. PPV: positive predictive value; NPV: negative predictive value; MPC: Matthews correlation coefficient.

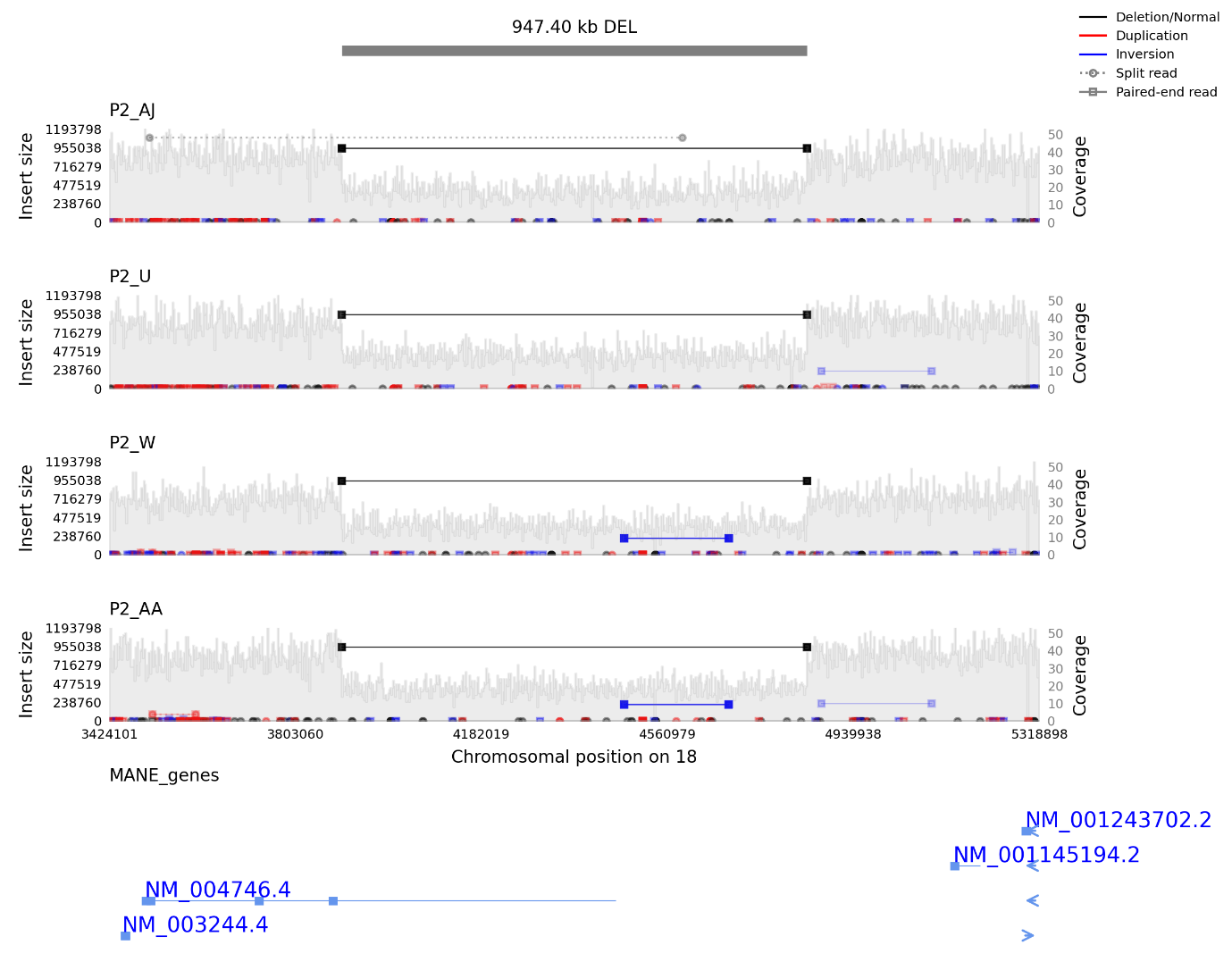

Supplementary Figure 4: A visualisation of the sequencing reads surrounding the *DLGAP1* deletion for the four carriers using samplot (Belyeu et al., 2021). The dark grey strip at the top indicates the CNV region. The pale grey bars indicate the depth of coverage (measured from the right-side y-axis). The horizontal lines represent paired-end or split reads, and their insert size (measured from the left-side y-axis). MANE transcripts are shown at the bottom of the plot, with coding exons represented by the blue boxes. *DLGAP1* is represented by transcript ID NM_004746.4.

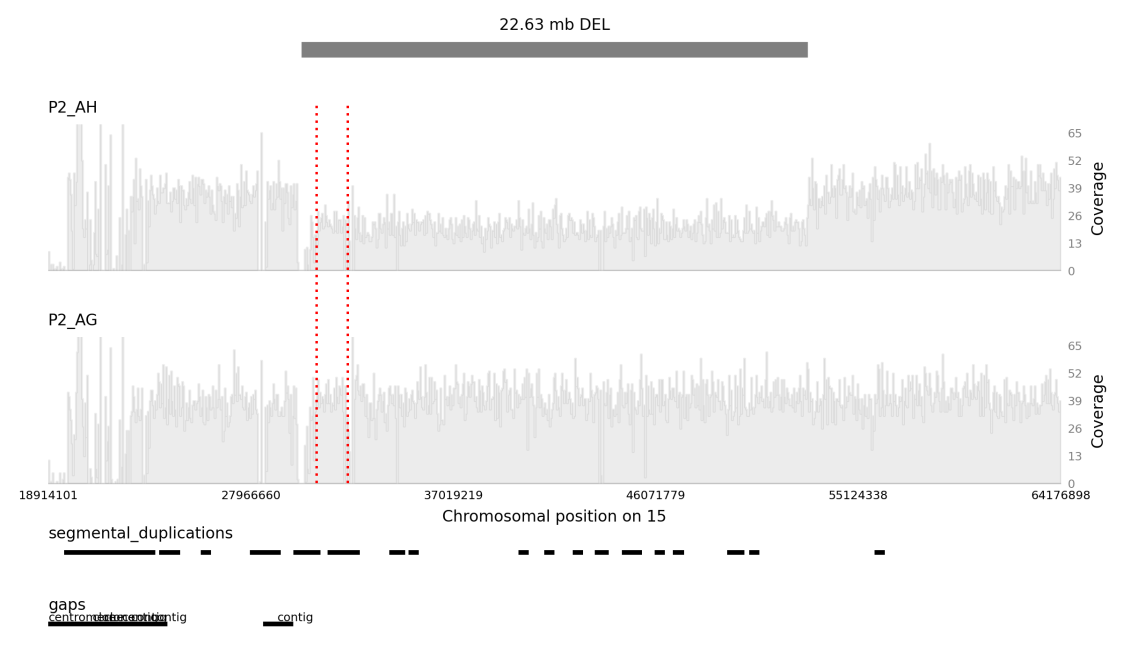

Supplementary Figure 5: A visualisation of the sequencing reads surrounding the 15q13.3 deletion using samplot (Belyeu et al., 2021). The dark grey strip at the top indicates the large deletion boundaries and the red dashed lines indicate the 15q13.3 critical region. The pale grey bars indicate the depth of coverage (measured from the right-side y-axis). The upper plot is for the deletion carrier (individual AH), and the lower plot is for their child (individual AG) who is not a carrier. For readability due to the length of the CNV region, the paired-end and split read data have been removed. Segmental duplications and gaps in the GRCh38 assembly are shown at the bottom of the plot.

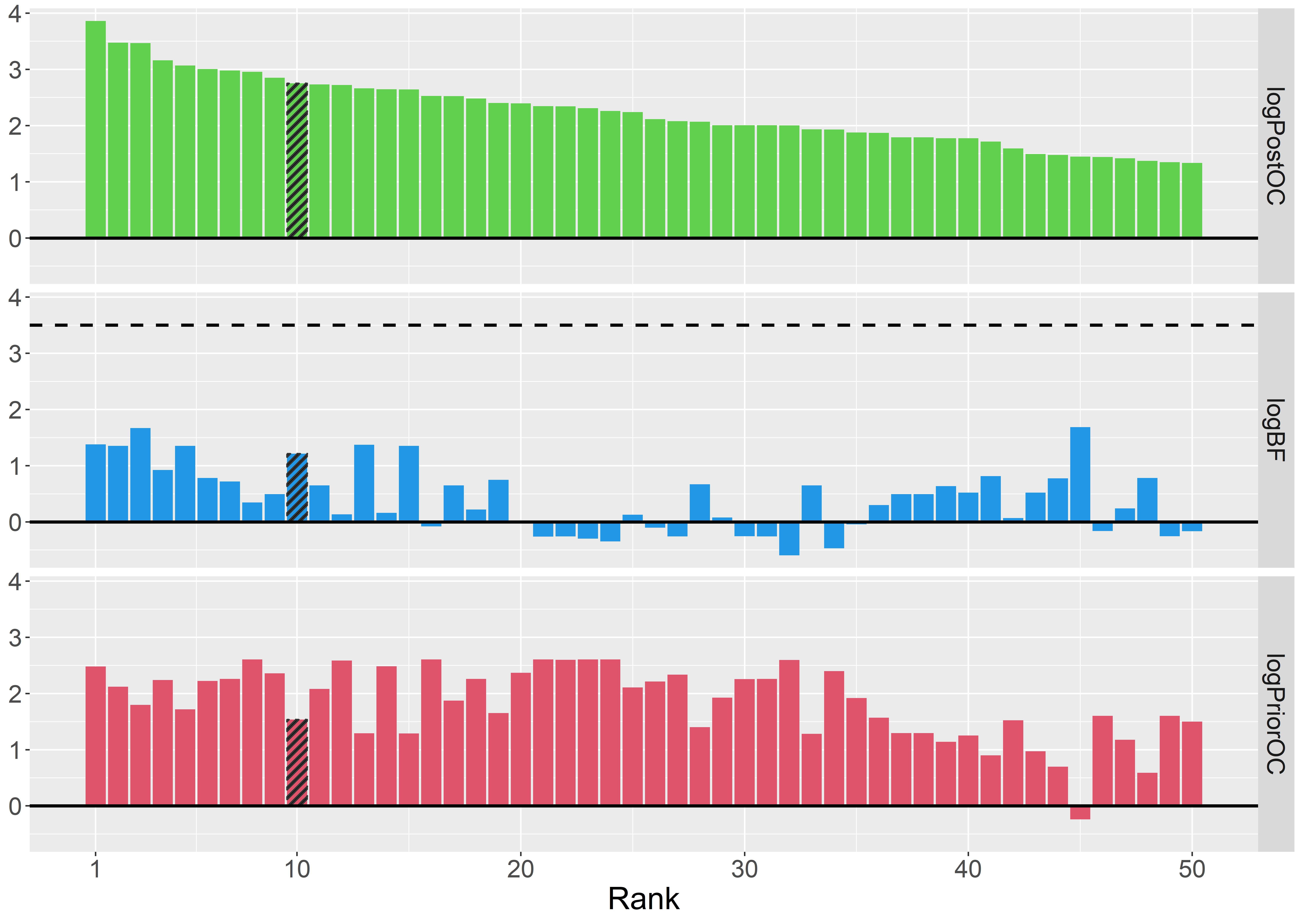

Supplementary Figure 6: BICEP output metrics for the top 50 ranked SNVs and indels in P1 with anxiety as the selected phenotype. The dashed line in the middle panel indicates the maximum logBF achievable in the pedigree. The shaded bars represent the *NPY5R* variant.

### 3 Supplementary Tables

| **Pedigree** | **Name** | **OCD** | **ADHD** | **CMVT** | **TS** | **Anxiety** |
| --- | --- | --- | --- | --- | --- | --- |
| P1 | 2 | Present | N/A | Absent | Absent | SAD, phobia |
|  | 4 | Present | N/A | Absent | Absent | SAD |
|  | 5 | Present | N/A | Absent | Absent | N/A |
|  | 9 | Present | N/A | Absent | Absent | GAD, SAD, PD |
|  | 11 | Present | N/A | Absent | Absent | GAD |
|  | 15 | Present | Present | Absent | Present | N/A |
|  | 18 | Probable | N/A | Absent | Absent | GAD, phobia |
|  | 20 | Present | N/A | Absent | Absent | GAD, SAD, phobia |
|  | 74 | Present | N/A | Absent | Absent | N/A |
|  | 143 | Probable | N/A | Absent | Absent | PD |
|  | 146 | Present | N/A | Absent | Absent | GAD |
|  | 149 | Present | N/A | Absent | Absent | N/A |
|  | 177 | Present | N/A | Absent | Absent | GAD, PD |
|  | 180 | Present | N/A | Probable | Absent | GAD |
|  | 197 | Present | N/A | Absent | Absent | GAD, SAD, PD |
| P2 | A | Probable | N/A | N/A | N/A | N/A |
|  | W | Absent | Absent | N/A | N/A | N/A |
|  | U | Present | N/A | N/A | N/A | N/A |
|  | AA | Present | Absent | N/A | N/A | N/A |
|  | AH | Present | Absent | N/A | N/A | N/A |
|  | AG | Present | Absent | N/A | N/A | N/A |
|  | J | Present | Probable | N/A | N/A | N/A |
|  | H | Present | Absent | N/A | N/A | N/A |
|  | S | Present | Present | N/A | N/A | N/A |
|  | AJ | Present | Absent | N/A | N/A | N/A |

Supplementary Table 1: Phenotypic information for the 25 individuals sequenced. N/A indicates no information available. OCD: obsessive compulsive disorder; ADHD: attention deficit/hyperactivity disorder; CMVT: chronic motor/vocal tic; TS: Tourette syndrome; SAD: separation anxiety disorder; PD: panic disorder; GAD: generalised anxiety disorder.

| **Chr** | **Start** | **End** | **Locus (gene)** | **Type** | **Overlap criteria** | **SCZ** | **ASD** | **OCD** | **TS** | **XD** |
| --- | --- | --- | --- | --- | --- | --- | --- | --- | --- | --- |
| chr1 | 145627235 | 146040039 | 1q21.1 | DEL | Size >50% of critical region | x |  |  |  |  |
| chr1 | 145627235 | 146040039 | 1q21.1 | DUP | Size >50% of critical region | x | x |  |  |  |
| chr1 | 147063850 | 147910423 | 1q21.1-21.2 | DEL | Size >50% of critical region | x |  |  |  |  |
| chr1 | 147063850 | 147910423 | 1q21.1-21.2 | DUP | Size >50% of critical region |  | x |  |  | x |
| chr1 | 245749346 | 246507279 | 1q44 (*SMYD3*) | DUP | Whole gene | x |  |  |  |  |
| chr2 | 49918502 | 51032132 | 2p16.3 (*NRXN1*) | DEL | Exonic deletions | x | x |  | x | x |
| chr3 | 1093023 | 1404217 | 3p26.3 (*CNTN6*) | DUP | Whole gene |  |  |  | x |  |
| chr3 | 195993296 | 197627955 | 3q29 | DEL | Size >50% of critical region | x | x |  |  | x |
| chr7 | 73330912 | 74728554 | 7q11.23, WBS | DUP | Size >50% of critical region | x | x |  |  |  |
| chr8 | 737627 | 1708476 | 8p23.3 (*DLGAP2*) | DEL | Exonic deletions |  | x |  |  |  |
| chr8 | 271671 | 4352618 | 8p23.3-23.2 (*DLGAP2;CSMD1*) | DEL | Exonic deletions |  | x |  |  |  |
| chr9 | 116423111 | 117415057 | 9q32 (*ASTN2*) | DEL | Exonic deletions |  |  |  |  | x |
| chr15 | 22778537 | 23067754 | 15q11.2 | DEL | Size >50% of critical region | x |  |  |  |  |
| chr15 | 22778537 | 23067754 | 15q11.2 | DUP | Size >50% of critical region |  | x |  |  |  |
| chr15 | 22999177 | 28099315 | 15q11-q13, AS/PWS | DUP | Full critical region, ~4Mbp | x | x |  |  |  |
| chr15 | 30788442 | 32170575 | 15q13.3 | DEL | Size >50% of critical region | x |  |  |  | x |
| chr15 | 30788442 | 32170575 | 15q13.3 | DUP | Size >50% of critical region |  | x |  |  |  |
| chr16 | 8933494 | 9163303 | 16p13.2 (*USP7;HAPSTR1*) | DUP | Whole gene | x |  |  |  | x |
| chr16 | 8892096 | 8963906 | 16p13.2 (*USP7*) | DUP | Whole gene |  | x |  |  | x |
| chr16 | 9091643 | 9121635 | 16p13.2 (*HAPSTR1*) | DUP | Whole gene | x |  |  |  | x |
| chr16 | 15417798 | 16199832 | 16p13.11 | DEL | Size >50% of critical region |  |  | ? |  | x |
| chr16 | 15417798 | 16199832 | 16p13.11 | DUP | Size >50% of critical region | x |  |  |  |  |
| chr16 | 21955548 | 22412377 | 16p12.2, BP2-BP3 | DEL | Size >50% of critical region | x |  |  |  | x |
| chr16 | 28811875 | 29035462 | 16p11.2, distal | DEL | Size >50% of critical region | x |  |  |  | x |
| chr16 | 29639519 | 30189452 | 16p11.2, proximal | DEL | Size >50% of critical region |  | x |  |  | x |
| chr16 | 29639519 | 30189452 | 16p11.2, proximal | DUP | Size >50% of critical region | x | x |  |  |  |
| chr22 | 19036286 | 21064168 | 22q11.2 | DEL | Size >50% of critical region | x |  |  |  | x |
| chr22 | 19036286 | 21064168 | 22q11.2 | DUP | Size >50% of critical region |  | x |  |  |  |
| chr22 | 50674408 | 50733212 | 22q13.3 (*SHANK3*) | DEL | At least 1Mbp, including *SHANK3* |  | x |  |  | x |

Supplementary Table 2: A list of rare deletion (DEL) and duplications (DUP) implicated in schizophrenia (SCZ) (Marshall et al., 2017; Rees et al., 2014), autism (ASD) (Sanders et al., 2015), obsessive-compulsive disorder (OCD) (McGrath et al., 2014), Tourette syndrome (TS) (Huang et al., 2017), and a cross-disorder analysis (XD) of psychiatric traits (Shanta et al., 2025). In the phenotype columns, ‘x’ indicates a significant association and ‘?’ indicates a nominal association. WBS: Williams-Beuren syndrome.; AS: Angelman syndrome; PWS: Prader-Willi syndrome.

| **Description** | **P1** | **P2**  **Founder E** | **P2**  **Founder E** | **P2**  **Founder A** |
| --- | --- | --- | --- | --- |
| Rank | 2 | 1 | 3 | 9 |
| Coordinates (GRCh38) | chr4:163351538:C:G | chr18:3897800-4845199 | chr16:1767576:G:A | chr3:47867332:T:C |
| Gene | *NPY5R* | *DLGAP1* | *MAPK8IP3* | *MAP4* |
| Variant type | Missense variant | Deletion | Missense variant | Missense variant |
| Amino acid change | Pro422Arg | N/A | Ala1084Thr | Ile2139Val |
| logPriorOC | 1.41 | 0.83 | 0.29 | 0.33 |
| logBF | 2.72 | 1.13 | 1.13 | 0.68 |
| logPostOC | 4.13 | 1.96 | 1.43 | 1.02 |
| Allele frequency | $3.1\times{10}^{-4}$ | $0\times{10}^{0}$ | $4.1\times{10}^{-6}$ | $1.5\times{10}^{-3}$ |
| FATHMM | -5.15 | N/A | 1.48 | -3.70 |
| MPC | 0.81 | N/A | 1.58 | 0.40 |
| PolyPhen2 HDIV | 1.00 | N/A | 1.00 | 0.99 |
| REVEL | 0.96 | N/A | 0.49 | 0.76 |
| SIFT | 0.00 | N/A | 0.00 | 0.01 |
| loeuf_sumRecip | N/A | N/A | N/A | N/A |
| CADD-SV | N/A | 12.97 | N/A | N/A |

Supplementary Table 3: Details of the rare variants with the most compelling evidence from BICEP or found in known risk genes. Included is the gnomAD v4 allele frequencies and the deleteriousness metrics used in the prior model. The amino acid changes are relative to the MANE transcript.

| **Description** | **P1** | | **P2**  **Founder A** | | **P2**  **Founder E** | |
| --- | --- | --- | --- | --- | --- | --- |
|  | **DEL** | **DUP** | **DEL** | **DUP** | **DEL** | **DUP** |
| Called by PECAN | 3,321 | 192 | 3,163 | 180 | 3,163 | 180 |
| Remove CNVs not carried by branch founder | 2,504 | 113 | 2,610 | 118 | 2,076 | 86 |
| Carried by at least 2 individuals | 2,504 | 113 | 1,671 | 39 | 1,190 | 29 |
| Gene disrupting | 840 | 39 | 530 | 16 | 368 | 11 |
| Length at least 30kbp | 21 | 10 | 11 | 4 | 8 | 1 |

Supplementary Table 4: Number of deletions (DEL) and duplications (DUP) retained at each stage of quality control filtering.

### 4 References

Belyeu, J. R., Chowdhury, M., Brown, J., Pedersen, B. S., Cormier, M. J., Quinlan, A. R., & Layer, R. M. (2021). Samplot: a platform for structural variant visual validation and automated filtering. *Genome Biol, 22*(1), 161. doi:10.1186/s13059-021-02380-5

Haeussler, M., Zweig, A. S., Tyner, C., Speir, M. L., Rosenbloom, K. R., Raney, B. J., . . . Kent, W. J. (2019). The UCSC Genome Browser database: 2019 update. *Nucleic Acids Res, 47*(D1), D853-d858. doi:10.1093/nar/gky1095

Huang, A. Y., Yu, D., Davis, L. K., Sul, J. H., Tsetsos, F., Ramensky, V., . . . Coppola, G. (2017). Rare Copy Number Variants in NRXN1 and CNTN6 Increase Risk for Tourette Syndrome. *Neuron, 94*(6), 1101-1111.e1107. doi:10.1016/j.neuron.2017.06.010

Marshall, C. R., Howrigan, D. P., Merico, D., Thiruvahindrapuram, B., Wu, W., Greer, D. S., . . . Sebat, J. (2017). Contribution of copy number variants to schizophrenia from a genome-wide study of 41,321 subjects. *Nat Genet, 49*(1), 27-35. doi:10.1038/ng.3725

McGrath, L. M., Yu, D., Marshall, C., Davis, L. K., Thiruvahindrapuram, B., Li, B., . . . Scharf, J. M. (2014). Copy number variation in obsessive-compulsive disorder and tourette syndrome: a cross-disorder study. *J Am Acad Child Adolesc Psychiatry, 53*(8), 910-919. doi:10.1016/j.jaac.2014.04.022

Ormond, C., Ryan, N. M., Cap, M., Byerley, W., Corvin, A., & Heron, E. A. (2024). BICEP: Bayesian inference for rare genomic variant causality evaluation in pedigrees. *Brief Bioinform, 26*(1). doi:10.1093/bib/bbae624

Ormond, C., Ryan, N. M., Corvin, A., & Heron, E. A. (2021). Converting single nucleotide variants between genome builds: from cautionary tale to solution. *Brief Bioinform, 22*(5). doi:10.1093/bib/bbab069

Pedersen, B. S., & Quinlan, A. R. (2017). Who's Who? Detecting and Resolving Sample Anomalies in Human DNA Sequencing Studies with Peddy. *Am J Hum Genet, 100*(3), 406-413. doi:10.1016/j.ajhg.2017.01.017

Purcell, S., Neale, B., Todd-Brown, K., Thomas, L., Ferreira, M. A., Bender, D., . . . Sham, P. C. (2007). PLINK: a tool set for whole-genome association and population-based linkage analyses. *Am J Hum Genet, 81*(3), 559-575. doi:10.1086/519795

Rees, E., Walters, J. T., Georgieva, L., Isles, A. R., Chambert, K. D., Richards, A. L., . . . Kirov, G. (2014). Analysis of copy number variations at 15 schizophrenia-associated loci. *Br J Psychiatry, 204*(2), 108-114. doi:10.1192/bjp.bp.113.131052

Sanders, S. J., He, X., Willsey, A. J., Ercan-Sencicek, A. G., Samocha, K. E., Cicek, A. E., . . . State, M. W. (2015). Insights into Autism Spectrum Disorder Genomic Architecture and Biology from 71 Risk Loci. *Neuron, 87*(6), 1215-1233. doi:10.1016/j.neuron.2015.09.016

Shanta, O., Klein, M., Sacks, M., MacDonald, J. R., Maihofer, A., Ahangari, M., . . . Sebat, J. (2025). A cross-disorder analysis of CNVs finds novel loci and dose-dependent relationships of genes to psychiatric traits. *medRxiv*. doi:10.1101/2025.07.11.25331310
